# Observational methods for COVID-19 vaccine effectiveness research: a trial emulation and empirical evaluation

**DOI:** 10.1101/2022.11.09.22282065

**Authors:** Martí Català, Edward Burn, Trishna Rathod-Mistry, Junqing Xie, Antonella Delmestri, Daniel Prieto-Alhambra, Annika M. Jödicke

## Abstract

Despite much research on the topic, little work has been done comparing the use of methods to control for confounding in the estimation of COVID-19 vaccine effectiveness in routinely collected medical record data. We conducted a trial emulation study to replicate the ChAdOx1 (Oxford/AstraZeneca) and BNT162b2 (BioNTech/Pfizer) COVID-19 phase 3 efficacy studies. We conducted a cohort study including individuals aged 75+ from UK CPRD AURUM (N = 916,128) in early 2021. Three different methods were assessed: Overlap weighting, inverse probability treatment weighting, and propensity score matching. All three methods successfully replicated the findings from both phase 3 trials, and overlap weighting performed best in terms of confounding, systematic error, and precision. Despite lack of trial data beyond 3 weeks, we found that even 1 dose of BNT162b2 was effective against SARS-CoV-2 infection for up to 12 weeks before a second dose was administered. These results support the UK Joint Committee on Vaccination and Immunisation modelling and related UK vaccination strategies implemented in early 2021.

**Key messages:**

- Real world evidence generated using weighting (overlapping weights and inverse probability of treatment weights) and propensity score matching: all methods successfully replicate the findings of Phase 3 trials for COVID-19 vaccine effectiveness.
- Overlap weighting provides the least biased estimates in our study and should be considered amongst the most suitable methods for future COVID-19 vaccine effectiveness research.
- Despite a lack of trial data, our findings suggest that first-dose BNT162b2 provides effective protection against SARS-COV-2 infection for up to 12 weeks, in line with UK’s Joint Committee on Vaccination and Immunisation modelling and subsequent vaccination strategies.

## Introduction

Following the start of the COVID-19 vaccination programs, routinely collected data are being widely used to evaluate the effectiveness and safety of COVID-19 vaccines(1-4) and boosters(5, 6). However, careful consideration of how-to best account for confounding is required when comparing vaccinated and unvaccinated people: Aside from people’s individual characteristics, particularly age and co-morbidities associated with higher risk of severe COVID-19, population-level confounder such as the location, i.e. level of community transmission, and vaccination period are among the most important confounders. However, the latter were not always adequately considered in many of the previously conducted vaccine effectiveness studies. Differences in methods for adjustment for confounders as well as choice of study design, inclusion criteria and calendar time can have substantial impact on the findings and their interpretation as highlighted in recent observational studies(7, 8) assessing the comparative effectiveness of ChAdOx1 (Oxford/AstraZeneca) and BNT162b2 (BioNTech/Pfizer) using routinely-collected data from UK: a 28% (95% CI, 12-42) decreased risk for infection after first dose vaccination with BNT162b2(8) vs. no differences in infection incidence between the two COVID-19 vaccines (7). Another challenge when studying vaccine effectiveness is the handling of the immediate time after the first vaccine dose. Randomised trials (9, 10) showed no protective vaccine effect in the first 2 weeks, with the vaccine-induced immunity still building up. However, while some observational studies could replicate these findings (1), others already observed protective effects in the early days (11, 12), indicating the presence of residual confounding. However, some studies and even trials omitted this vulnerable time from their main analyses (10) while others did not (9), which makes it difficult to compare results across studies.

While several methods were used in previous studies, a rigorous assessment of their ability to resolve confounding has not yet been completed(13). Our study provides an empirical evaluation of the comparative performance of different weighting and matching methods to minimise confounding in the study of COVID-19 vaccine effectiveness: overlap weighting(14), inverse probability of treatment weights(15), and propensity score with exact geographic and index date matching. To do this, we measured confounding based on imbalances between vaccinated vs unvaccinated subjects in terms of all recorded variables in the patient’s records. Additionally, we used negative control outcomes to identify systematic error due to unobserved confounders. Lastly, we conducted target trial emulations to compare our findings to those from the BNT162b2(9) and ChAdOx1(10) phase 3 randomised controlled trials. As no trials assessed the effect of BNT162b2 between 3 and 12 weeks following the first dose, we estimated vaccine effectiveness based on primary care data and compared our results to the models the Joint Committee on Vaccination and Immunisation (JCVI) used to inform the vaccination campaign in the UK in early 2021(16).

## Methods

### Study type, setting, and data source

A cohort study was conducted using UK NHS primary care data from the Clinical Practice Research Datalink (CPRD) AURUM, mapped to the Observational Medical Outcomes Partnership (OMOP) Common Data Model (CDM)(17).

### Study population

All people aged ≥ 75 years, who were not previously infected with or vaccinated against SARS-CoV-2 and were registered in CPRD AURUM for ≥180 days before study start (04/01/2021) were eligible for inclusion. Subsequently, individuals were assigned to the vaccinated (VC) or unvaccinated (UV) cohort, based on whether they were vaccinated against COVID-19 between 04/01/2021 and 28/01/2021 (both dates included). Two different vaccinations were recorded at that time in England(18): BNT162b2 (BioNTech/Pfizer) and ChAdOx1 (Oxford/AstraZeneca). We therefore constructed three different vaccinated cohorts: any type of vaccination cohort (VC), ChAdOx1 vaccinated cohort (AZ), and BNT162b2 vaccinated cohort (PF). Index date for individuals in the vaccinated cohort was their vaccination date. All individuals who had a record of both vaccines at the index date were excluded. Assignment of the index date for unvaccinated people depended on the method used to account for confounding, i.e. matching or weighting. For matching, the index date of the matched counterpart was used; whereas for PS weighting, index dates for unvaccinated people were randomly assigned following the distribution of index dates in the vaccinated cohort, as depicted in Figure 1. In supplementary material Figure S1 distribution of index dates for vaccine type effectiveness are shown. After assignment of the respective index dates, individuals with a recording of COVID-19 infection before or on index date and individuals vaccinated before index date were excluded.

**Figure 1.**
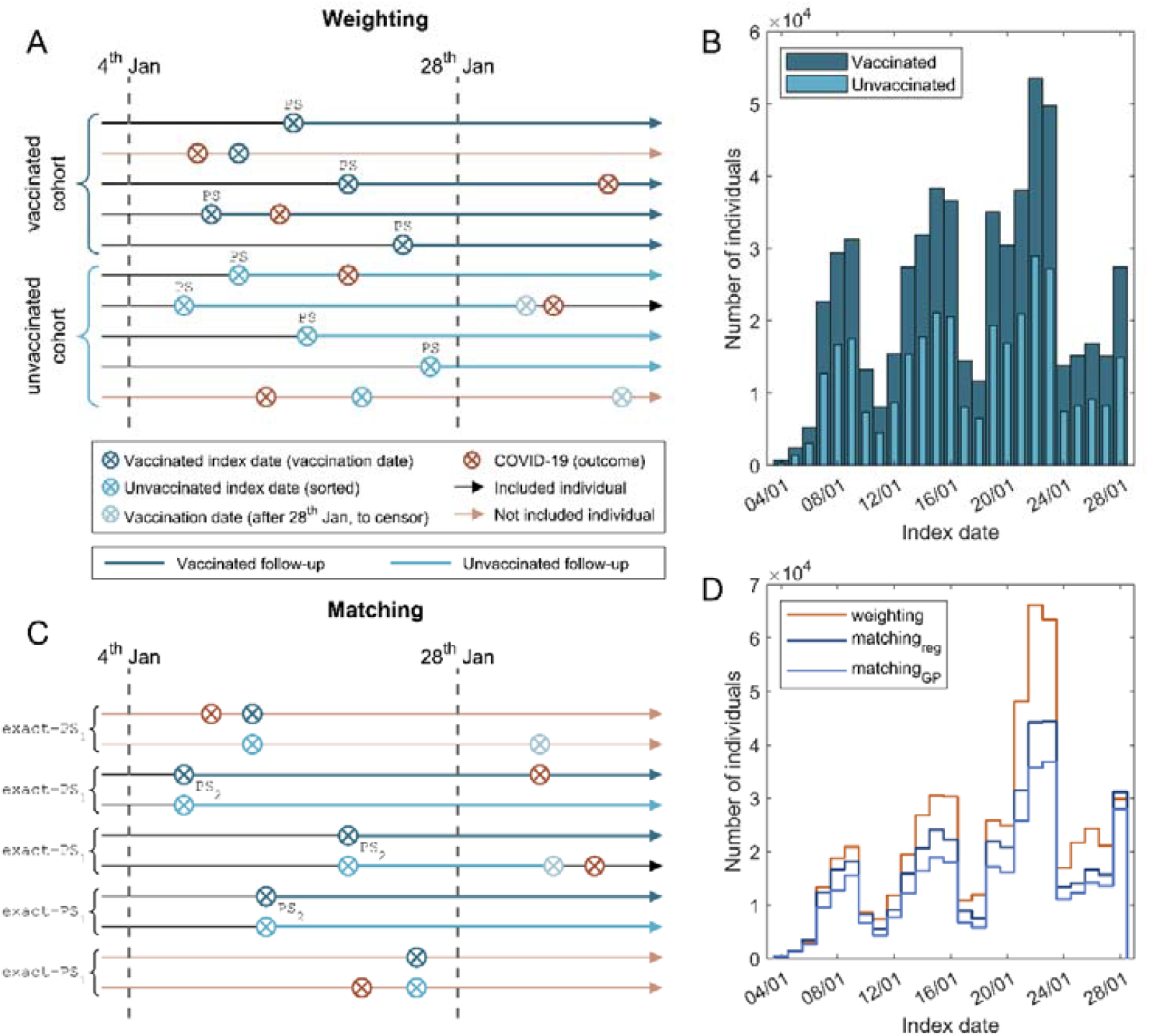
Index date and follow-up in vaccinated and unvaccinated cohorts. (A) Diagram depicting index date and follow up for both vaccinated and unvaccinated cohorts in weighting analyses; exclusion based on COVID-19 infection status. Follow-up is represented by a thicker/wider solid arrow. (B) Distribution of index dates for vaccinated (dark blue) and unvaccinated cohorts (light blue). (C) Diagram depicting index date and follow up for both vaccinated and unvaccinated participants in matching analyses; exclusion based on COVID-19 infection status. Follow-up period is represented by a thicker line. (D) Distribution of index dates for weighting and different matching analyses.

### Exposure/s of interest

Each of the three identified vaccinated cohorts (VC, PF, AZ) were compared with an unvaccinated cohort (UV) to study vaccine effectiveness after weighting or matching.

### Follow-up

Individuals were censored when: they received another vaccination dose (vaccinated cohorts) or received the first vaccination dose (unvaccinated); left the CPRD AURUM database; or they died.

### Primary Outcomes

Two clinical outcomes were ascertained, in line with primary outcomes in the target phase 3 trials for the vaccines: (i) COVID-19 PCR, defined by a positive COVID-19 PCR test; and (ii) COVID-19 PCR/diagnosis, defined by either a COVID-19 diagnosis or a positive COVID-19 test. Definitions of the clinical outcomes can be found in supplementary material Tables S1-3.

### Control Outcomes

#### PCR testing

We used PCR testing (e.g. performed COVID-19 PCR test regardless of test result) as a control outcome and proxy for diagnostic effort. We would expect this to be non-differential with respect to vaccination.

#### Negative control outcomes (NCO)

Additionally, a total of 43 NCO were pre-specified based on previous methodological research on vaccine safety(19) and refined after review by two clinical epidemiologists (DPA and AMJ). NCO are outcomes not causally associated with the exposure of interest, here COVID-19 vaccine/s exposure. Detail on the 43 utilised NCO is provided in the supplementary material Table S4.

### Statistical analyses

#### Propensity Score

Large-scale Propensity Scores (PS) were used to minimise confounding. PS represent the probability of exposure based on a participant’s baseline characteristics. Covariates to be included in the PS equation were extracted, including condition occurrences for three different time windows (1 to 30 days, 31 to 180 days, and 181 days to any time prior index date), and drug exposures for 2 time periods (1 to 30 days, 31 to 180 days prior index date). Subsequently, all covariates with a frequency >0.5% were included in a lasso regression, which was used to identify relevant confounders to be included in the large-scale PS. In addition, the following variables were forced into the PS model as they were known to be associated with vaccination in the UK: location (region identifier; 9 different regions; or General Practice (GP) surgery identifier; 1357 different GP); age (5-year bands and as a continuous variable using a polynomial to account for non-linearity); prior observation years; number of GP visits, and number of previous COVID-19 PCR tests during each of the three periods (1 to 30-day, 31 to 180-day and 181 days to any time before index date).

#### PS Weighting and Matching

Regional vaccination, testing, and COVID-19 incidence rates(20) on index date were forced into the PS. Index date was also forced into the PS to ensure that the balance is maintained. PS were computed using a logistic regression model, with 3 different representations of location: without location (PS_base_), location defined as region (PS_reg_) or de-identified GP surgery (PS_GP_). We used two different weighting methods: Inverse Probability of Treatment Weighting (IPTW), with trimming over 0.95 and above 0.05; and Overlap Weighting (OW)(21).

As for *Matching*, we used 1:1 PS nearest neighbours matching with exact matching on age band, gender, and location (region or GP) with caliper width 0.2 SD on the PS as calculated on their index date (PS_1_). Matched unvaccinated individuals were assigned the same index date as their vaccinated pair. PS were computed again on the new index date after matching (PS_2_) to ensure that the matching was still balanced after the index date assignment.

The following metrics were used to assess the performance of the different methods:

1. Covariate imbalance as a proxy of measured confounding was assessed by calculating standardized mean differences (SMD) between compared cohorts after weighting or matching.
2. Minimum detectable relative risk (MDRR) was computed for a 95% confidence interval, power of 0.8, a 10-day cumulative incidence of 0.0067, and 10 days of following time.
3. Association between vaccination status and Negative Control Outcomes (NCO)(22) was estimated using Cox proportional hazard regression to detect unmeasured confounding.
4. Association between vaccination status and COVID-19 PCR/diagnosis in the first 10 days following vaccination was estimated using Cox proportional hazard regression to detect unmeasured confounding through comparison with results from vaccination trials(9).

#### Empirical calibration

Hazard ratios calculated for (1) PCR testing, (2) COVID-19 PCR and (3) COVID-19 PCR/diagnosis were calibrated with the result of the NCO analysis as described in Schuemie et al.(23).

#### Target trial emulations

We aimed to reproduce the effectiveness observed at 3 weeks after vaccination in the BNT162b2 and in the ChAdOx1 phase 3 trials(9, 10) and the estimated effectiveness at 12 weeks for BNT162b2. For the AstraZeneca trial emulation, individuals who tested positive or were censored before the 24^th^ day were eliminated from the analysis in line with the trial’s statistical analysis plan and protocol(10). We deemed a target trial as successfully replicated when the confidence intervals from the trial and observational data overlapped. It is worth noting that the 12-week trial estimate of vaccine effectiveness for BNT162b2 was based on an extrapolation by the UK JCVI committee, and did not report confidence intervals(16). Estimates from observational data were obtained using the method yielding lowest confounding (lower SMD values and least number of statistically significant NCO) and better statistical power (narrower MDRR).

### Patient and Public Involvement

A patient representative was involved in the design of the project.

## Results

### Study population and characteristics

Following application of inclusion criteria, 583,813 vaccinated individuals (348,275 PF and 235,538 AZ); and 332,315 unvaccinated participants were identified (Figure 2). OW weighting methods retained the largest sample size, with a total of 905,418 participants included for analysis. IPTW led to the inclusion of the second largest number of participants, with N=903,147 for IPTW-PS_base_, N=902,958 for IPTW-PS_reg_, and N=856,636 for IPTW-PS_GP_. Due to the exclusion of unmatched people, matching methods led to a substantially reduced sample size: N=459,000 for matching_reg_; N=369,310 for matching_GP_. Population flowcharts stratified by vaccine type are available in supplementary material Figures S2-3.

**Figure 2.**
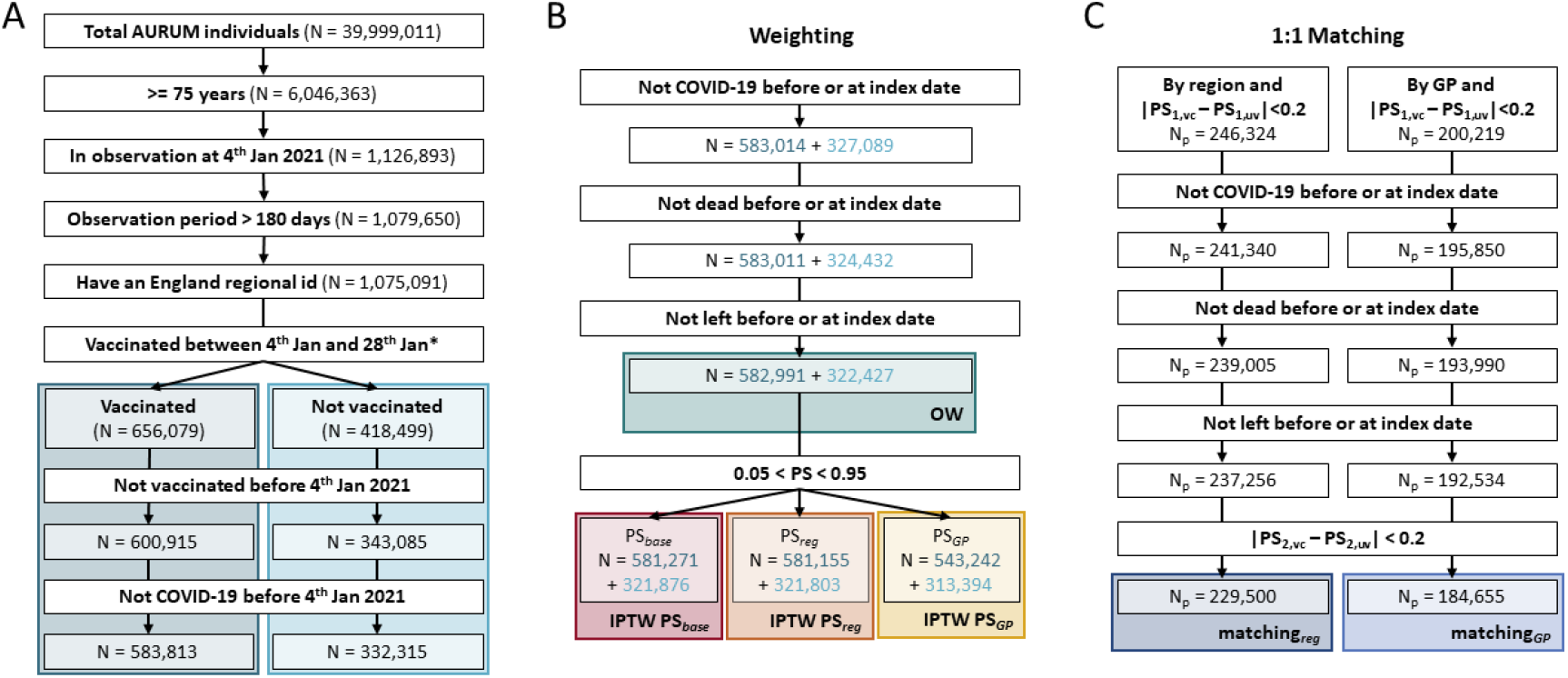
Cohorts building flowchart in any type of vaccination and unvaccinated comparison. **(A)** Flowchart to build the any type of vaccination (VC) and unvaccinated (UV) initial cohorts. **(B)** Flowchart to build the different weighting cohorts, the start point of these cohorts is the end of panel A. Dark blue numbers are for vaccinated cohort and light for unvaccinated. **(C)** Flowchart to build the different matching cohorts, the start point of these cohorts is the end of panel A. PS_1_ and PS_2_ are the propensity scores (PS) computed at the start and index date, respectively. *At this step individuals with a record of both ChAdOx1 and BNT162b2 vaccines at the index date were excluded.

Baseline characteristics before weighting/matching are shown in Table 1. Relevant differences (SMD>0.1) between vaccinated and unvaccinated cohorts included age, location (region and GP practice) and number of GP visits in the previous 180 days. Additionally, differences were noted for comorbidities heart disease, hypertensive disorder, malignant neoplastic disease, and renal impairment. Supplementary material Tables S5-7 provide detail on a total of 23 imbalanced confounders with SMD > 0.1 in the comparison of vaccinated vs unvaccinated, 25 in AZ vs unvaccinated, and 27 in PF vs unvaccinated cohorts.

**Table 1.** Baseline differences between unvaccinated cohort and the vaccinated cohorts before weighting/matching. The different covariates included in this table are computed at the start date (4_th_ Jan) for each individual. Age and prior observation time are measured in years. Region and General Practice (GP) surgery are the location identifiers. GP visits and number of PCR tests are only recorded for the last 180 days. Comorbidities are recorded for any time prior. Standardised mean differences (SMD) are computed compared to the unvaccinated cohort.

|  | unvaccinated | vaccinated |  |  |  |  |  |
| --- | --- | --- | --- | --- | --- | --- | --- |
|  | (N=332,315) | Any type<br>(N=583,813) | SMD | AstraZeneca<br>(N=235,538) | SMD | Pfizer<br>(N=348,275) | SMD |
| **Age** (median [IQR]) | 78 [76-83] | 81 [78-86] | 0.285 | 80 [77-85] | 0.208 | 82 [78-86] | 0.341 |
| **Age groups** N (%) |  |  | 0.509 |  | 0.310 |  | 0.655 |
| 75 to 79 years | 207,710 (62.5%) | 224,388 (38.4%) |  | 111,630 (47.4%) |  | 112,758 (32.4%) |  |
| 80 to 84 years | 58,693 (17.7%) | 183,171 (31.4%) |  | 61,821 (26.2%) |  | 121,350 (34.8%) |  |
| 85 to 89 years | 36,015 (10.8%) | 110,762 (19.0%) |  | 34,455 (14.6%) |  | 76,307 (21.9%) |  |
| 90 to 94 years | 21,070 (6.3%) | 49,449 (8.5%) |  | 19,299 (8.2%) |  | 30,150 (8.7%) |  |
| 95 to 99 years | 7,332 (2.2%) | 13,887 (2.4%) |  | 7,005 (3.0%) |  | 6,882 (2.0%) |  |
| >99 years | 1,495 (0.4%) | 2,156 (0.4%) |  | 1,328 (0.6%) |  | 828 (0.2%) |  |
| **Gender** Male N (%) | 147,234 (44.3%) | 252,761 (43.3%) | 0.020 | 99,268 (42.1%) | 0.044 | 153,493 (44.1%) | 0.005 |
| **Region** N (%) |  |  | 0.141 |  | 0.204 |  | 0.148 |
| North East | 10,595 (3.2%) | 18,834 (3.2%) |  | 8,026 (3.4%) |  | 10,808 (3.1%) |  |
| North West | 54,543 (16.4%) | 119,038 (20.4%) |  | 47,890 (20.3%) |  | 71,148 (20.4%) |  |
| Yorkshire And The Humber | 10,580 (3.2%) | 21,895 (3.8%) |  | 11,221 (4.8%) |  | 10,674 (3.1%) |  |
| East Midlands | 8,079 (2.4%) | 10,205 (1.7%) |  | 4,350 (1.8%) |  | 5,855 (1.7%) |  |
| West Midlands | 61,028 (18.4%) | 106,134 (18.2%) |  | 41,550 (17.6%) |  | 64,584 (18.5%) |  |
| East of England | 12,591 (3.8%) | 28,832 (4.9%) |  | 12,785 (5.4%) |  | 16,047 (4.6%) |  |
| South West | 48,825 (14.7%) | 79,330 (13.6%) |  | 36,369 (15.4%) |  | 42,961 (12.3%) |  |
| South East-Central | 80,764 (24.3%) | 125,969 (21.6%) |  | 51,225 (21.7%) |  | 74,744 (21.5%) |  |
| London | 45,310 (13.6%) | 73,576 (12.6%) |  | 22,122 (9.4%) |  | 51,454 (14.8%) |  |
| **GP surgery** |  |  | 0.725 |  | 0.815 |  | 0.974 |
| Different GP surgeries | 1355 | 1356 |  | 1352 |  | 1343 |  |
| Individuals per GP surgery (median [IQR]) | 169 [87-319] | 324 [151-593] |  | 114 [46-227] |  | 173 [71-348] |  |
| **Prior Observation time** (median [IQR]) | 23.1 [9.6-34.9] | 25.3 [11.4-37.1] | 0.065 | 24.2 [9.8-36.3] | 0.033 | 25.9 [12.6-37.7] | 0.086 |
| **GP visits** (median [IQR]) 180 days | 9 [4-17] | 22 [12-36] | 0.624 | 22 [12-36] | 0.634 | 22 [12-34] | 0.617 |
| **Number PCR tests** (mean±STD) 180 days | 0.1±0.6 | 0.3±1.7 | 0.141 | 0.6±2.2 | 0.211 | 0.2±1.2 | 0.072 |
| **Comorbidities** N (%) |  |  |  |  |  |  |  |
| Asthma | 38,193 (11.5%) | 70,385 (12.1%) | 0.017 | 28,329 (12.0%) | 0.017 | 42,056 (12.1%) | 0.018 |
| Autoimmune disease | 13,920 (4.2%) | 26,281 (4.5%) | 0.015 | 10,893 (4.6%) | 0.021 | 15,388 (4.4%) | 0.011 |
| COPD | 28,627 (8.6%) | 52,176 (8.9%) | 0.011 | 21,619 (9.2%) | 0.020 | 30,557 (8.8%) | 0.006 |
| Dementia | 18,340 (5.5%) | 37,944 (6.5%) | 0.041 | 20,147 (8.6%) | 0.119 | 17,797 (5.1%) | 0.018 |
| Diabetes mellitus | 64,137 (19.3%) | 112,685 (19.3%) | 0.000 | 45,403 (19.3%) | 0.001 | 67,282 (19.3%) | 0.000 |
| Heart disease | 101,742 (30.6%) | 211,270 (36.2%) | 0.118 | 83,292 (35.4%) | 0.101 | 127,978 (36.7%) | 0.130 |
| Hypertensive disorder | 185,562 (55.8%) | 355,249 (60.8%) | 0.102 | 141,100 (59.9%) | 0.082 | 214,149 (61.5%) | 0.115 |
| Malignant neoplastic disease | 76,186 (22.9%) | 160,017 (27.4%) | 0.103 | 61,967 (26.3%) | 0.079 | 98,050 (28.2%) | 0.120 |
| Renal impairment | 80,367 (24.2%) | 169,014 (29.0%) | 0.108 | 66,110 (28.1%) | 0.088 | 102,904 (29.5%) | 0.121 |

### Comparison of performance of PS methods

Minimum detectable relative risks (MDRR) were computed for each of the comparison and methods as a proxy for statistical power and are reported in Table 2. As expected, the study was well powered due to large sample sizes, and MDRR was closer to one (better power) for weighting (e.g. MDRR 0.93 to 1.08 for IPTW in the ‘any vaccine’ analysis), and slightly further away for the one (worse power) for matching (e.g. MDRR 0.86 to 1.17 for matching_gp_ in the ‘any vaccine’ analysis). This relates to the exclusion of unmatched participants in the latter (see Figure 2). Standardized mean differences (SMD) and the number (%) of NCO associated with exposure are also reported in Table 2, as a proxy for measured/observed and unmeasured confounding, respectively. Both were highest in the unweighted analysis, and lowest in matching and OW compared to IPTW. Accounting for GP practice minimised confounding further: e.g. maximum SMD went from 0.79 in IPTW PS_base_ to 0.15 in IPTW PS_gp_. Overall, OW PS_gp_ and matching_gp_ were the best methods in terms of observed and unobserved confounding minimisation, measured by SMD and NCO respectively.

**Table 2.**
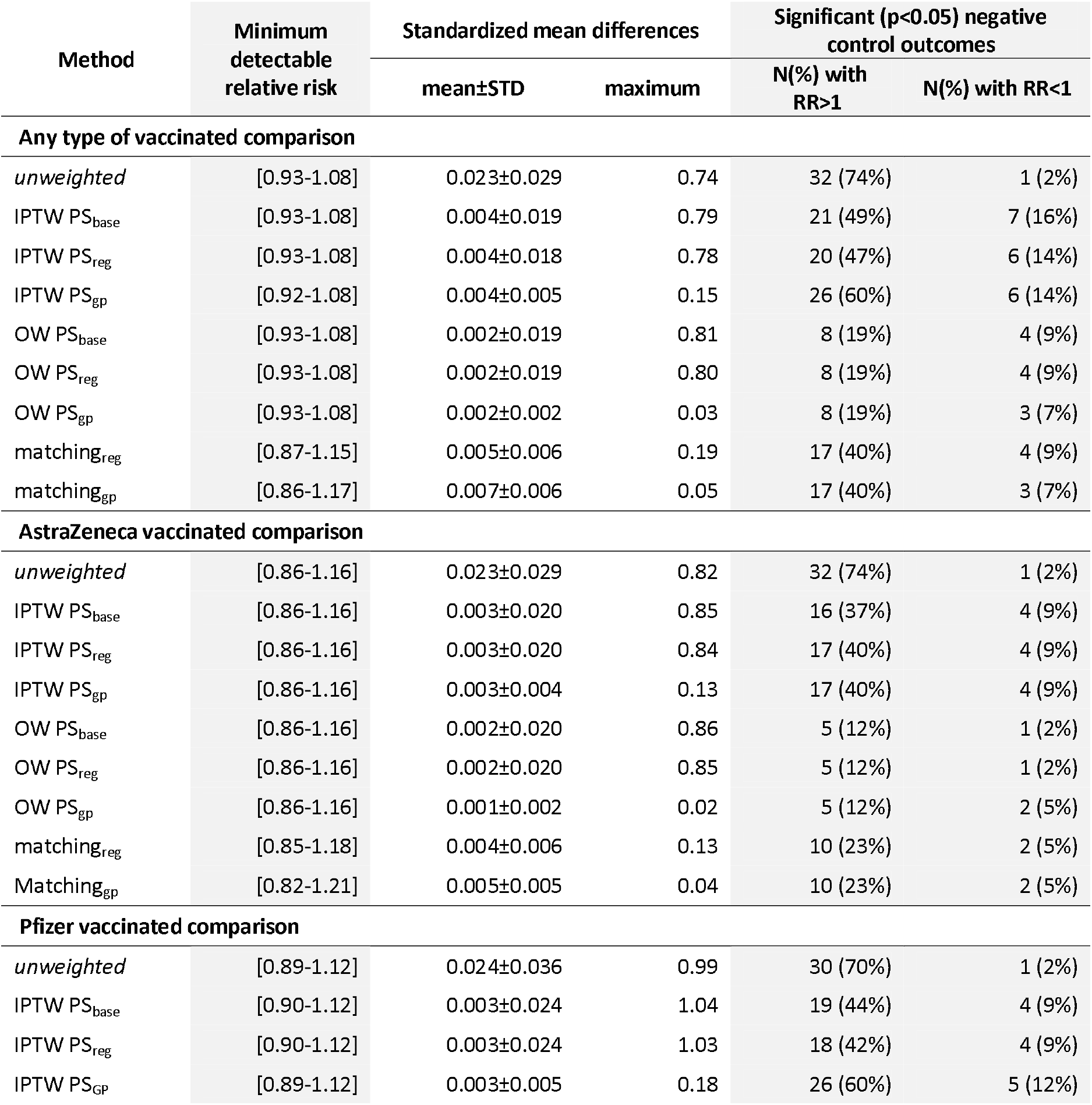
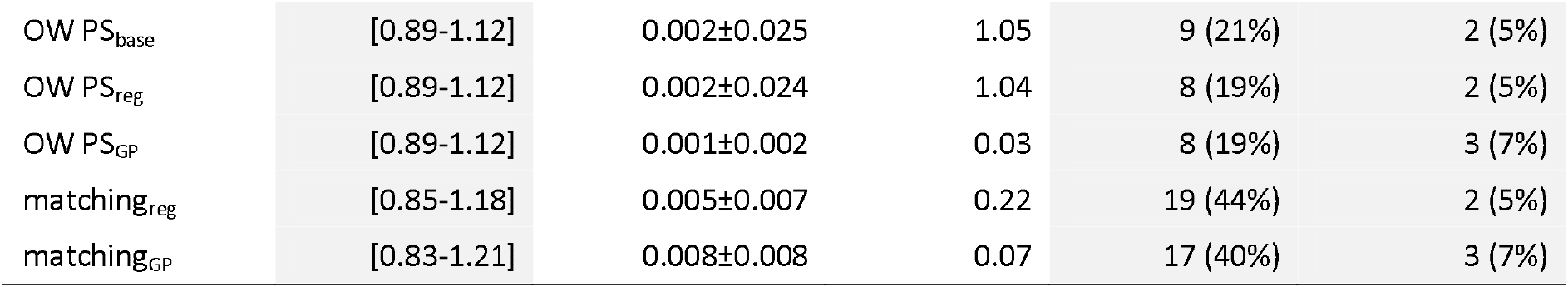
Minimum detectable relative risk (range); standardized mean differences (mean, standard deviation, and maximum for all the identified covariates); and number of significative negative control outcomes (positive and negative; regarding if they are positively or negatively associated with the exposure).

Figure 3 depicts SMD values for the weighted cohorts. Despite PS weighting, the GP practice covariate was only balanced using OW with PS_GP_. IPTW did not yield sufficient balance for GP practice in any of the estimated PS, as noted by SMD>0.1. Similar findings were seen for region, which was not balanced in any of the weighting methods unless location was included in the model. Matching at regional level did not yield sufficient balance for GP. In fact, only matching_gp_ and OW PS_gp_ were able to minimize GP unbalance. OW performed better than IPTW and matching in terms of overall covariate balance, with lower minimum SMD for all covariates (Table 2). Supplementary material Figures S4 and S5 show the SMD balancing for the Pfizer and AstraZeneca vs unvaccinated comparisons.

**Figure 3.**
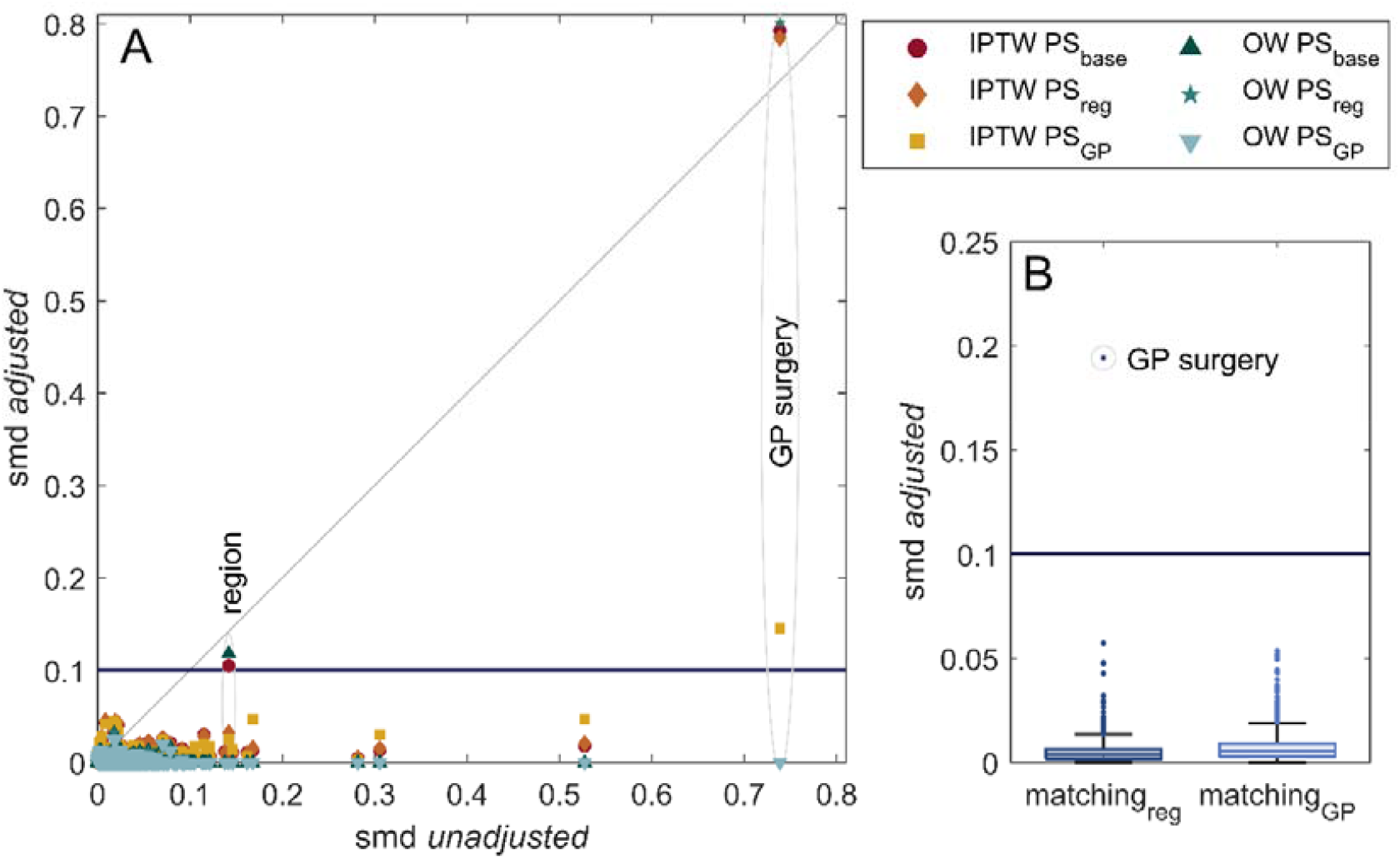
Standardized mean differences (SMD) for the different methods in any type of vaccination and unvaccinated comparison. (A) Scatter plot to compare before (unadjusted) and after (adjusted) weighting. Region and GP surgery are the only covariates that remain unbalanced after many of the weightings. (B) Boxplot for the covariates smd after matching. Only GP surgery is unbalanced after regional matching.

Figure 4 depicts the results of NCO analyses. In general, OW showed lower systematic error than IPTW and matching in most scenarios. Unweighted analyses show, as expected, clear evidence of one-sided systematic error, with many negative control outcomes being positively associated (RR>1) with vaccine exposure (Table 2). Supplementary material Figures S6 and S7 show the NCO results for the Pfizer and AstraZeneca vs unvaccinated comparisons.

**Figure 4.**
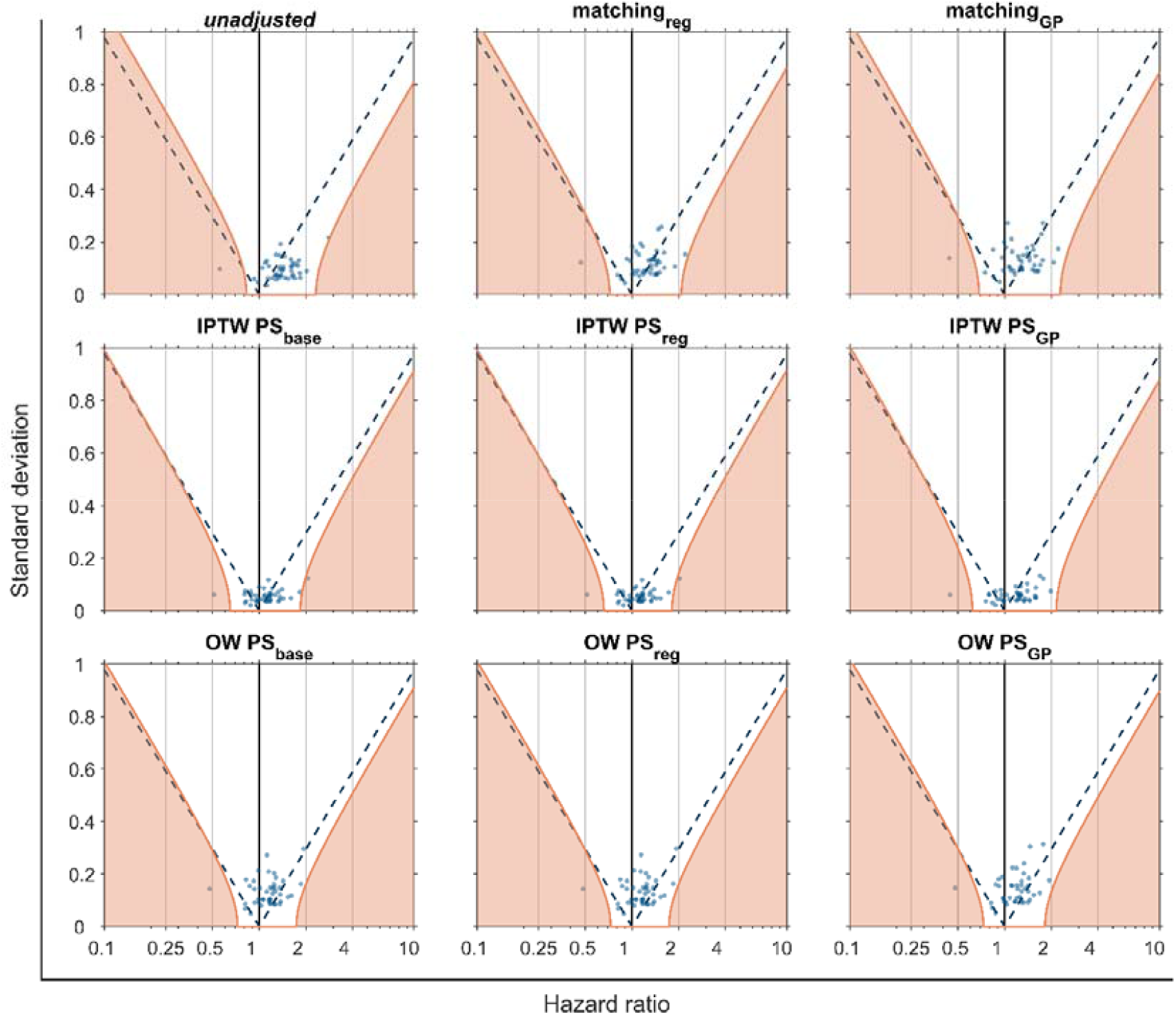
Negative control outcomes (NCO) hazard ratios and standard deviation. Each blue dot represents a different NCO. Purple dashed are the significative threshold for the NCO; they are positively correlated if they are on the right of the dashed line and negatively on the left. Orange lines mark significance thresholds after calibration.

In addition to the pre-specified NCO, Figure 5 shows the observed HR for the control outcome of PCR testing) and for the clinical outcomes (PCR+ and PCR+ or diagnosis) at day 10 after the first dose of vaccine. Although unweighted analyses led to a biased estimate (HR>1) for PCR testing, all weighting and adjustment methods resolved this and led to a calibrated HR including the expected null effect (HR=1). Unexpectedly, all the tested methods showed a protective effect against both clinical outcomes in the first 10 days after first-dose vaccination. The numerical values of Figure 5 can be observed in supplementary material Table S8. In supplementary material Figure S8 and Table S9 we can observe the same analysis than in Figure 5 without the 10 days censoring.

**Figure 5.**
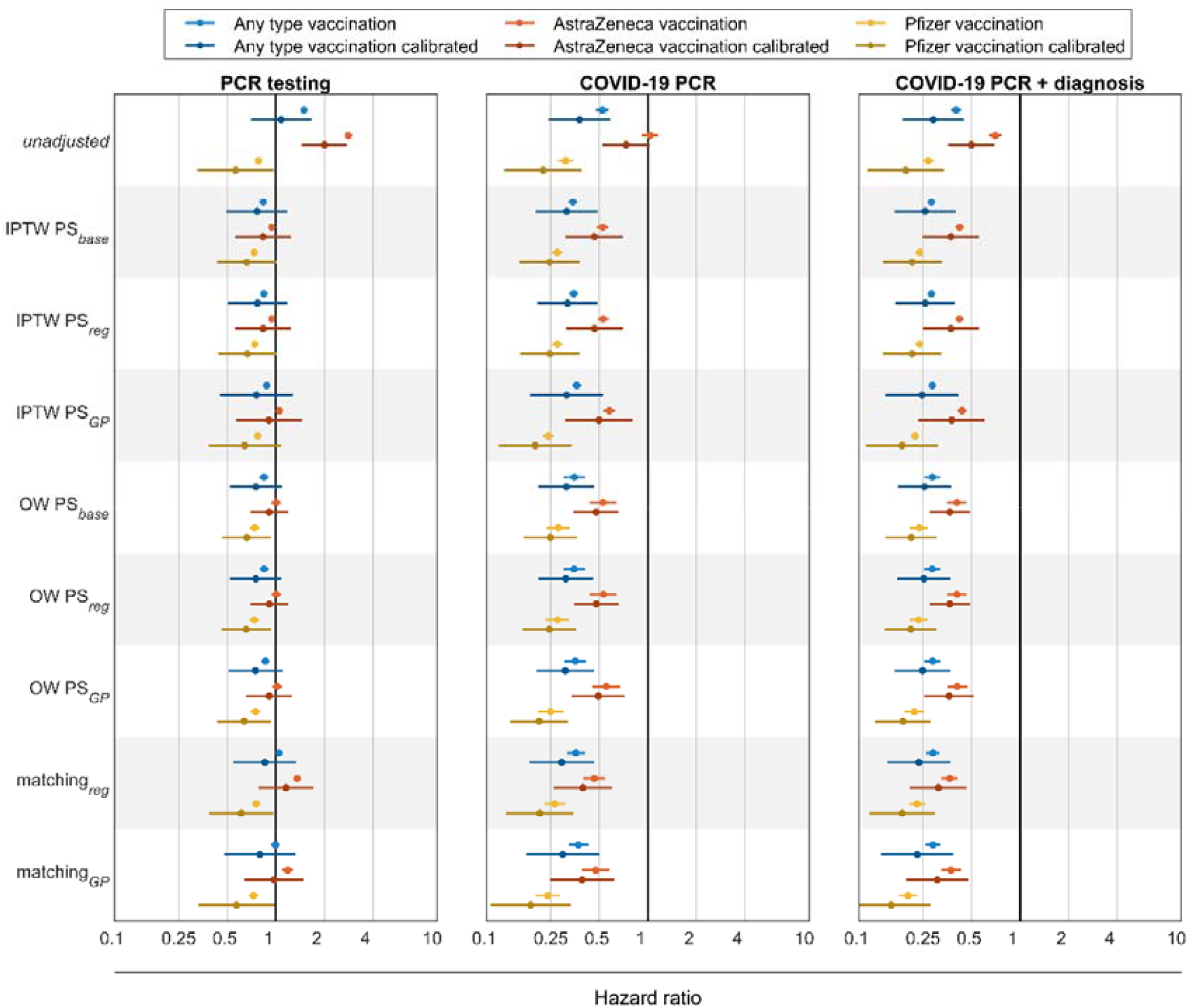
Hazard ratio for the control outcomes censoring at day 10. Each dot is the hazard ratio (HR) for a different adjustment computed with a Cox proportional hazards regression. Blue lines are for any type of vaccination compared to unvaccinated, red ones for AstraZeneca vaccinated compared to unvaccinated and yellow lines are for Pfizer vaccinated compared to unvaccinated. Darker lines are for calibrated hazard ratios. Vertical black line marks the HR = 1 threshold. In the left panel HR are for PCR testing, central and right panel for different COVID-19 definitions: only PCR positive and PCR positive or a diagnose, respectively.

The Kaplan-Meier plots using Overlapping Weights and PS_GP_ for the three studies comparisons can be observed in supplementary material Figure S9 (COVID-19 PCR + diagnosis) and Figure S10 (COVID-19 PCR).

### Target trial emulations

Figure 6 shows the results of the target trial emulations based on OW_gp_ analyses, as these yielded lowest bias and best statistical power. For ChAdOx1, we successfully replicated the results of vaccine effectiveness against COVID-19 PCR/diagnosis, with the calibrated estimate being closer to the trial result. As for BNT162b2, the observational estimates replicated both the 3- and the 12-week trial results only when calibrated results were used. Uncalibrated HRs overestimated effectiveness at week 3, and seemed to underestimate it at week 12 after first-dose vaccination.

**Figure 6.**
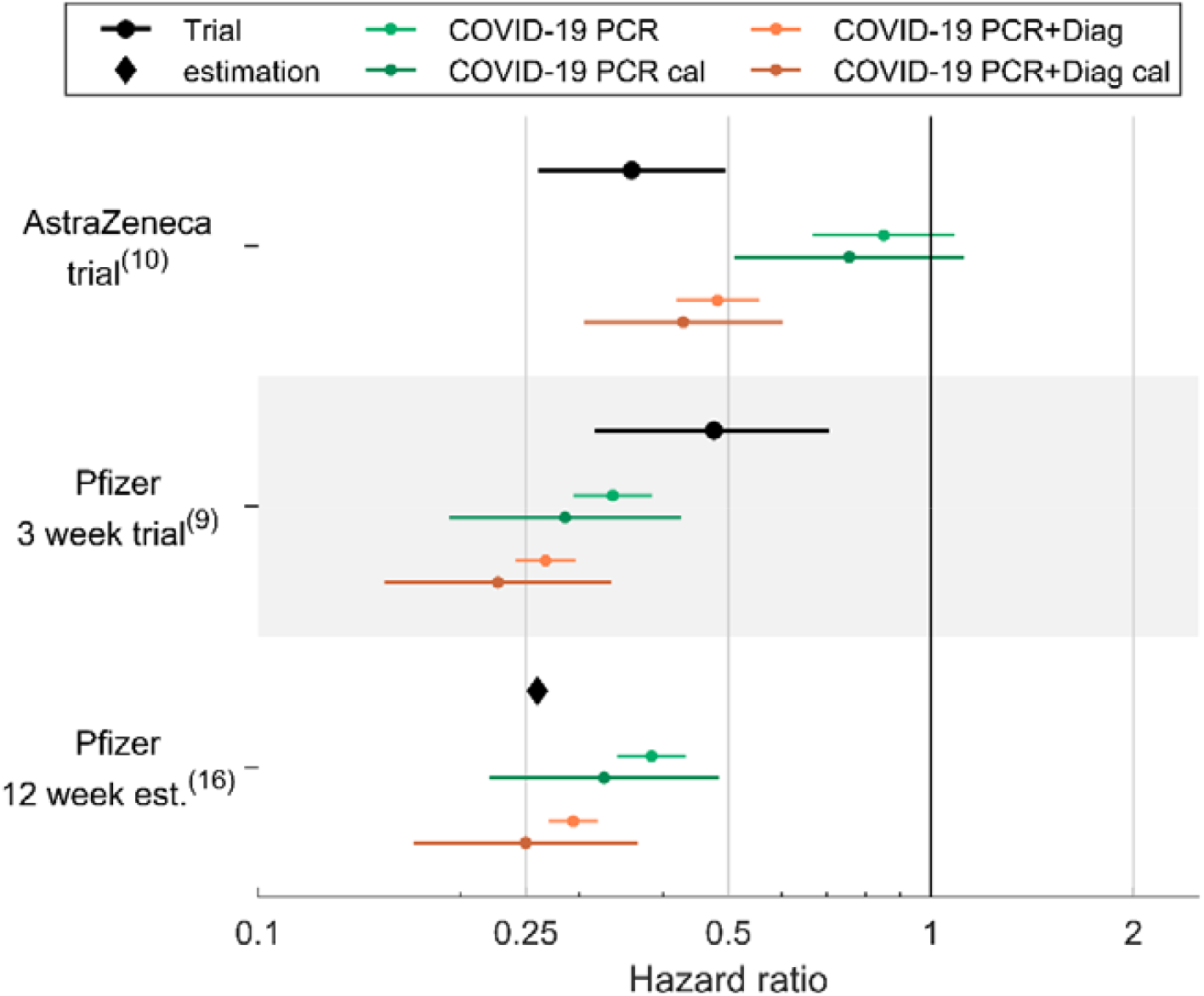
Trial emulation. Calibrated and uncalibrated relative risk for COVID-19 PCR and COVID-19 PCR and/or diagnose. For AstraZeneca trial individuals with a positive test within the first 21 days were eliminated.

## Discussion

In this large cohort study using data from UK primary care records, we assessed the effectiveness of the first dose of the widely used COVID-19 vaccines BNT162b2 and ChAdOx1 using overlap weighting. Our estimates of vaccine effectiveness are in line with trial estimates for the ChAdOx1 vaccine (57%) and BNT162b2 at 3 weeks (77%), as well as the JCVI estimates for BNT162b2 at 12 weeks (75%).

We chose overlap weighting for the target trial emulation, following an extensive investigation of the performance of different methods to account for confounding when studying COVID-19 vaccine effectiveness using observational data. Notably, merely including individual-level patient characteristics in the construction of PS did not guarantee the balance of population-level geographic variables, such as geographical region and GP, regardless of subsequent PS approaches of matching or weighting. Also, even if these geographic variables were incorporated into the propensity score, balance between groups was not sufficiently achieved using any of the studied methods, except for overlap weighting and matching. Additionally, negative control outcomes demonstrated a better resolution of uncontrolled confounding and systematic error with overlap weighting compared to other matching and weighting methods. Although no remarkable differences were seen in the final estimates of vaccine effectiveness based on this single database, the proposed theoretical advantages of overlap weighting(14, 21), including exact balance of every measured patient characteristics and good retainment of sample size, were supported by this empirical study. For the first time, we showed that overlap weighting overperforms other weighting and matching methods in terms of unmeasured confounding.

Importantly for COVID-19 vaccine effectiveness, overlap weighting was also the best performing method to minimise confounding at the population-level, including geographic location, which has important implications due to known differences in the spread of SARS-CoV-2 during the study period. Previous early vaccine effectiveness studies using observational data overlooked the adjustment of geographic information(24, 25). Our study, consistent with previous methodological literature, showed that geographic distributions between the vaccinated and unvaccinated individuals were markedly different(26), emphasizing the need for adjustment in vaccine effectiveness studies, given known differences in community transmission levels during the study period. In addition, the insufficient covariates’ balance after applying conventional methods highlighted the importance of the proposed diagnostics in observational studies of vaccine effectiveness. More importantly, our findings didn’t support the previous suspicion that less active health-seeking behaviour in unvaccinated people might confound the vaccine effectiveness observed in real-world data(12), as no association was found between vaccination and PCR testing in our study after the empirical calibration. The observed protective vaccine effects shortly after vaccination were in line with previous findings from observational data(11, 12), which might be attributable to differences in personal behaviour during the period immediately before and after vaccination, e.g. mask wearing, avoiding crowed indoor places/shielding, rescheduling vaccination if feeling unwell, which unfortunately was unlikely to be captured by electronic health records.

## Conclusions

Our study found vaccination with a first vaccine dose against COVID-19 associated with a 69% reduced risk of COVID-19 for both the ChAdOx1 and BNT162b2 vaccines. These data therefore confirm the estimation by JCVI that one dose of BNT162b2 provide protection beyond the 3-week period studied in pivotal trials. Secondly, we demonstrate that the studied propensity score weighting and matching methods can replicate pivotal trials and therefore provide reliable estimates of vaccine effectiveness. Further, PS-based overlap weights performed better than IPTW or matching in controlling for measured and unmeasured covariates while retaining sample size, and could be proposed as the preferred method for future vaccine effectiveness studies. Finally, our findings illustrate the need to incorporate patient location (e.g. GP practice or region of residence) and related variables (e.g. testing and transmission rates) to minimize community-as well as patient-level confounding in the study of COVID-19 vaccine effectiveness.

## Data Availability

Data were obtained from CPRD under the CPRD multi-study license held by the University of Oxford after Research Data Governance (RDG) approval. Direct data sharing is not allowed.

## Contributors and sources

MC led data analyses and contributed to analytical choices and study design. DPA and AMJ led the conceptualization of the study with contributions from EB and TRM. EB and TRM led the statistical analysis plan jointly with MC, DPA and AMJ. AD mapped and curated the data. MC, JX, DPA and AMJ wrote the first draft of the manuscript. All authors read, made contributions, and approved the last version of the manuscript. DPA and AMJ obtained the funding for this research.

## Ethics approval

The study protocol was approved through the CPRD’s Research Data Governance Process (Protocol No 21_000557).

## Funding

The project was supported by the National Institute for Health and Care Research (NIHR) Grant number COV-LT2-0006.

## Conflict of Interest

DPA receives funding from the UK National Institute for Health and Care Research (NIHR) in the form of a senior research fellowship and from the Oxford NIHR Biomedical Research Centre. His research group has received funding from the European Medicines Agency and Innovative Medicines Initiative. His research group has received research grant/s from Amgen, Chiesi-Taylor, GSK, Novartis, and UCB Biopharma. His department has also received advisory or consultancy fees from Amgen, Astellas, Astra Zeneca, Johnson and Johnson, and UCB Biopharma; and speaker fees from Amgen and UCB Biopharma. Janssen and Synapse Management Partners have supported training programmes organised by DPA’s department and open for external participants organized by his department outside the submitted work.

## Acknowledgements

This study is based in part on data from the Clinical Practice Research Datalink obtained under license from the UK Medicines and Healthcare products Regulatory Agency. The data are provided by patients and collected by the NHS as part of their care and support. The interpretation and conclusions contained in this study are those of the authors alone.⍰

## Supplementary material

### 1. Clinical definition and concept list

**Table S1.** PCR concept id clinical definition.

| Name | id | Class | Domain | Vocabulary |
| --- | --- | --- | --- | --- |
| Coronavirus nucleic acid detection | 44789510 | Procedure | Measurement | SNOMED |

**Table S2.**
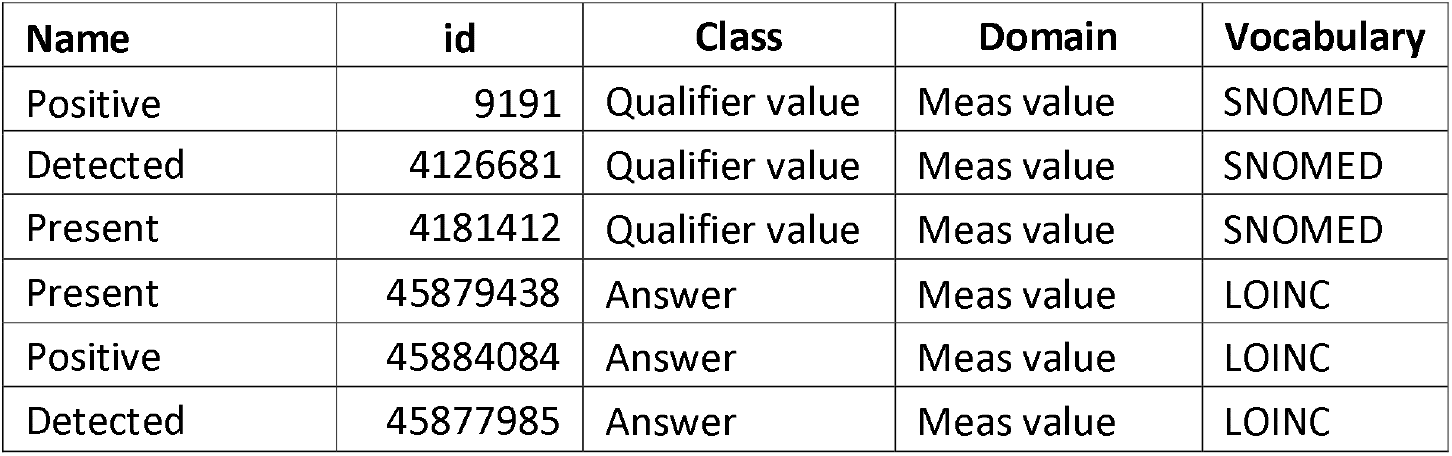
PCR results included in PCR+ clinical definition.

**Table S3.**
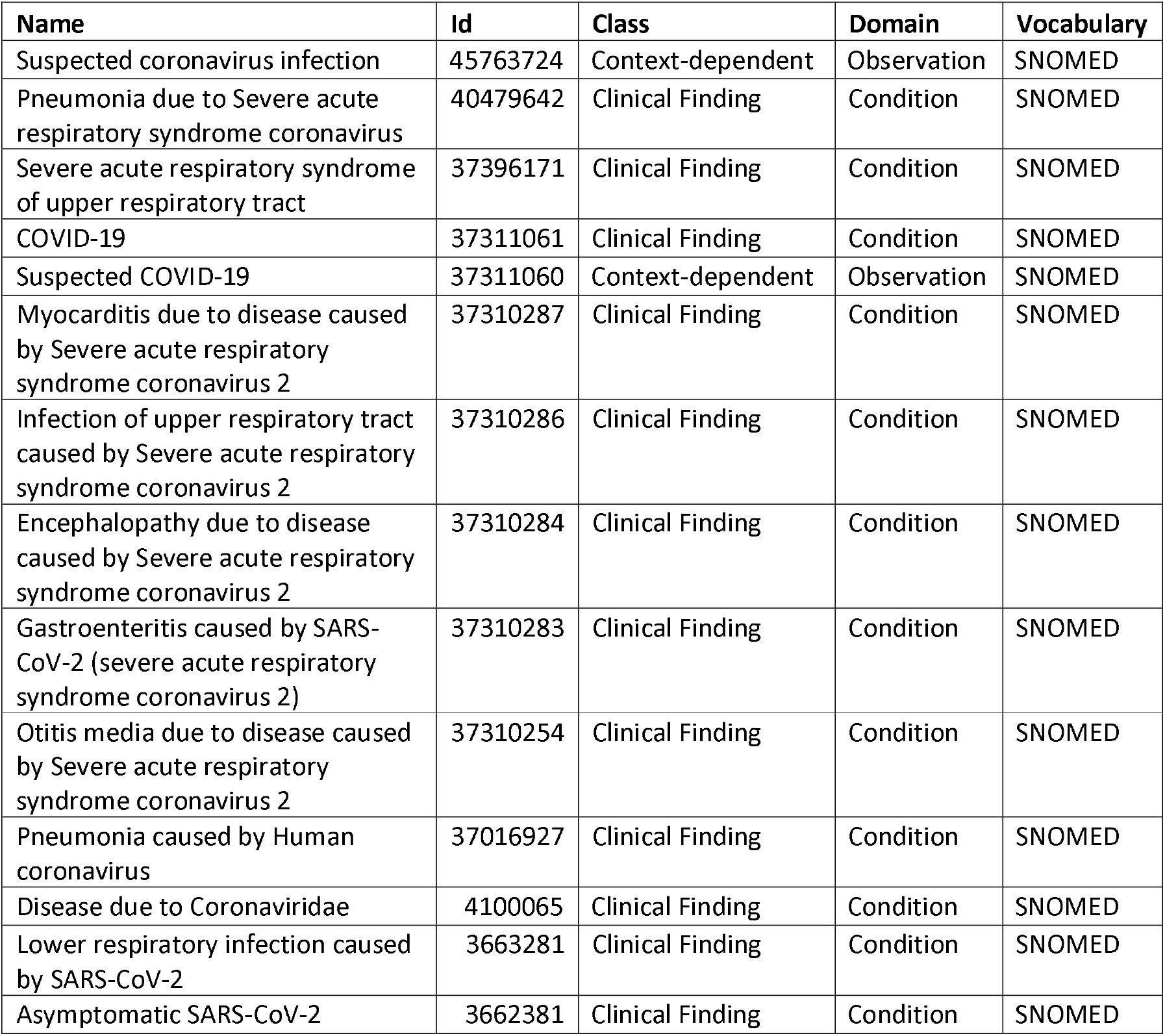
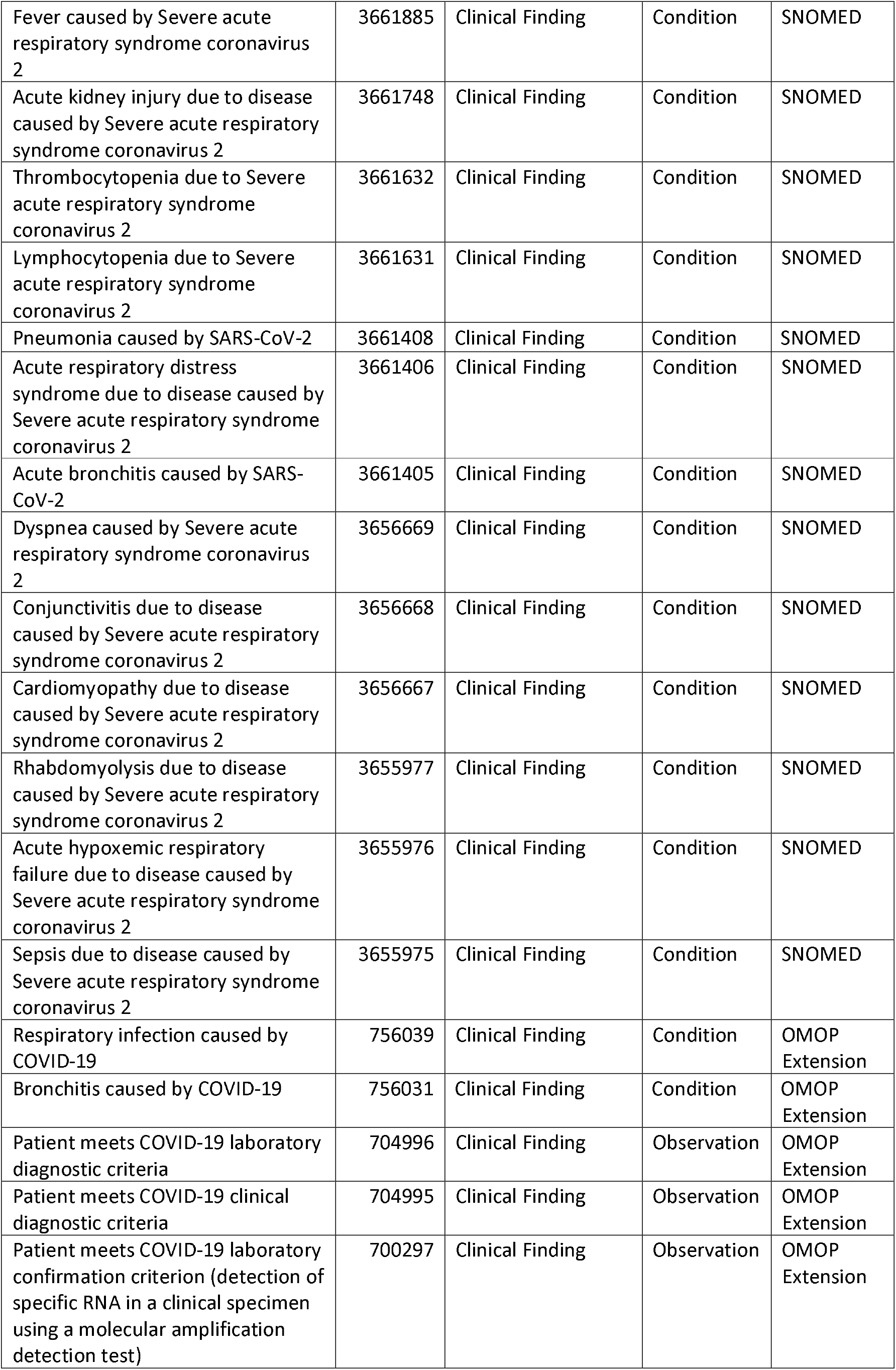
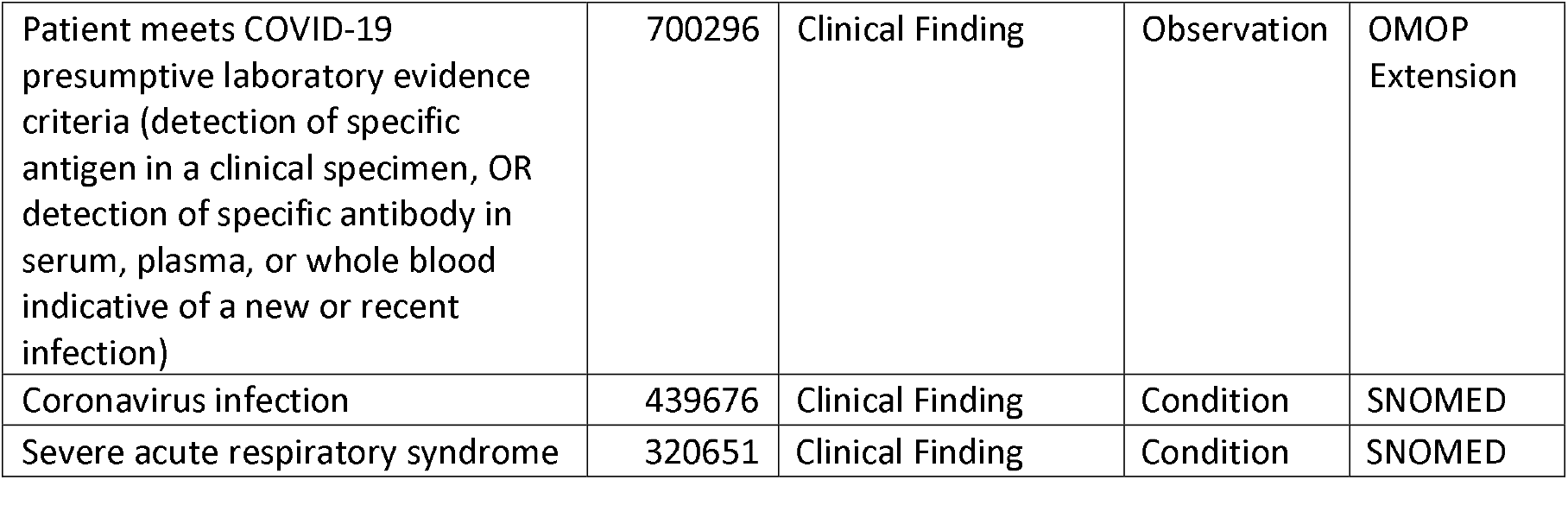
List of concept id included in COVID-19 clinical diagnosis definition.

| Name | Id | Class | Domain | Vocabulary |
| --- | --- | --- | --- | --- |
| Suspected coronavirus infection | 45763724 | Context-dependent | Observation | SNOMED |
| Pneumonia due to Severe acute respiratory syndrome coronavirus | 40479642 | Clinical Finding | Condition | SNOMED |
| Severe acute respiratory syndrome of upper respiratory tract | 37396171 | Clinical Finding | Condition | SNOMED |
| COVID-19 | 37311061 | Clinical Finding | Condition | SNOMED |
| Suspected COVID-19 | 37311060 | Context-dependent | Observation | SNOMED |
| Myocarditis due to disease caused by Severe acute respiratory syndrome coronavirus 2 | 37310287 | Clinical Finding | Condition | SNOMED |
| Infection of upper respiratory tract caused by Severe acute respiratory syndrome coronavirus 2 | 37310286 | Clinical Finding | Condition | SNOMED |
| Encephalopathy due to disease caused by Severe acute respiratory syndrome coronavirus 2 | 37310284 | Clinical Finding | Condition | SNOMED |
| Gastroenteritis caused by SARS-CoV-2 (severe acute respiratory syndrome coronavirus 2) | 37310283 | Clinical Finding | Condition | SNOMED |
| Otitis media due to disease caused by Severe acute respiratory syndrome coronavirus 2 | 37310254 | Clinical Finding | Condition | SNOMED |
| Pneumonia caused by Human coronavirus | 37016927 | Clinical Finding | Condition | SNOMED |
| Disease due to Coronaviridae | 4100065 | Clinical Finding | Condition | SNOMED |
| Lower respiratory infection caused by SARS-CoV-2 | 3663281 | Clinical Finding | Condition | SNOMED |
| Asymptomatic SARS-CoV-2 | 3662381 | Clinical Finding | Condition | SNOMED |
| Fever caused by Severe acute respiratory syndrome coronavirus 2 | 3661885 | Clinical Finding | Condition | SNOMED |
| Acute kidney injury due to disease caused by Severe acute respiratory syndrome coronavirus 2 | 3661748 | Clinical Finding | Condition | SNOMED |
| Thrombocytopenia due to Severe acute respiratory syndrome coronavirus 2 | 3661632 | Clinical Finding | Condition | SNOMED |
| Lymphocytopenia due to Severe acute respiratory syndrome coronavirus 2 | 3661631 | Clinical Finding | Condition | SNOMED |
| Pneumonia caused by SARS-CoV-2 | 3661408 | Clinical Finding | Condition | SNOMED |
| Acute respiratory distress syndrome due to disease caused by Severe acute respiratory syndrome coronavirus 2 | 3661406 | Clinical Finding | Condition | SNOMED |
| Acute bronchitis caused by SARS-CoV-2 | 3661405 | Clinical Finding | Condition | SNOMED |
| Dyspnea caused by Severe acute respiratory syndrome coronavirus 2 | 3656669 | Clinical Finding | Condition | SNOMED |
| Conjunctivitis due to disease caused by Severe acute respiratory syndrome coronavirus 2 | 3656668 | Clinical Finding | Condition | SNOMED |
| Cardiomyopathy due to disease caused by Severe acute respiratory syndrome coronavirus 2 | 3656667 | Clinical Finding | Condition | SNOMED |
| Rhabdomyolysis due to disease caused by Severe acute respiratory syndrome coronavirus 2 | 3655977 | Clinical Finding | Condition | SNOMED |
| Acute hypoxemic respiratory failure due to disease caused by Severe acute respiratory syndrome coronavirus 2 | 3655976 | Clinical Finding | Condition | SNOMED |
| Sepsis due to disease caused by Severe acute respiratory syndrome coronavirus 2 | 3655975 | Clinical Finding | Condition | SNOMED |
| Respiratory infection caused by COVID-19 | 756039 | Clinical Finding | Condition | OMOP Extension |
| Bronchitis caused by COVID-19 | 756031 | Clinical Finding | Condition | OMOP Extension |
| Patient meets COVID-19 laboratory diagnostic criteria | 704996 | Clinical Finding | Observation | OMOP Extension |
| Patient meets COVID-19 clinical diagnostic criteria | 704995 | Clinical Finding | Observation | OMOP Extension |
| Patient meets COVID-19 laboratory confirmation criterion (detection of specific RNA in a clinical specimen using a molecular amplification detection test) | 700297 | Clinical Finding | Observation | OMOP Extension |
| Patient meets COVID-19 presumptive laboratory evidence criteria (detection of specific antigen in a clinical specimen, OR detection of specific antibody in serum, plasma, or whole blood indicative of a new or recent infection) | 700296 | Clinical Finding | Observation | OMOP Extension |
| Coronavirus infection | 439676 | Clinical Finding | Condition | SNOMED |
| Severe acute respiratory syndrome | 320651 | Clinical Finding | Condition | SNOMED |

**Table S4.** Negative control outcomes concept id clinical definitions.

| Negative control outcome | id | Class | Domain | Vocabulary |
| --- | --- | --- | --- | --- |
| Acid reflux | 44783954 | Clinical Finding | Condition | SNOMED |
| Acquired hypothyroidism | 138384 | Clinical Finding | Condition | SNOMED |
| Actinic keratosis | 138825 | Clinical Finding | Condition | SNOMED |
| Acute conjunctivitis | 376707 | Clinical Finding | Condition | SNOMED |
| Age related macular degeneration | 374028 | Clinical Finding | Condition | SNOMED |
| Basal cell carcinoma of skin | 4112752 | Clinical Finding | Condition | SNOMED |
| Benign prostatic hyperplasia | 198803 | Clinical Finding | Condition | SNOMED |
| Bilateral cataracts | 4317977 | Clinical Finding | Condition | SNOMED |
| Blepharitis | 378425 | Clinical Finding | Condition | SNOMED |
| Cataract | 375545 | Clinical Finding | Condition | SNOMED |
| Cellulitis of lower limb | 42709838 | Clinical Finding | Condition | SNOMED |
| Constipation | 75860 | Clinical Finding | Condition | SNOMED |
| Dry eyes | 4036620 | Clinical Finding | Condition | SNOMED |
| Foot pain | 4169905 | Clinical Finding | Condition | SNOMED |
| Gallstone | 196456 | Clinical Finding | Condition | SNOMED |
| Glaucoma | 437541 | Clinical Finding | Condition | SNOMED |
| Hearing difficulty | 4038030 | Clinical Finding | Condition | SNOMED |
| Hearing loss | 377889 | Clinical Finding | Condition | SNOMED |
| Hemorrhoids | 195562 | Clinical Finding | Condition | SNOMED |
| Hypothyroidism | 140673 | Clinical Finding | Condition | SNOMED |
| Impacted cerumen | 374375 | Clinical Finding | Condition | SNOMED |
| Inguinal hernia | 4288544 | Clinical Finding | Condition | SNOMED |
| Intraocular pressure left eye | 4217260 | Clinical Finding | Condition | SNOMED |
| Iron deficiency anemia | 436659 | Clinical Finding | Condition | SNOMED |
| Laceration - injury | 443419 | Clinical Finding | Condition | SNOMED |
| Laceration of lower leg | 4155040 | Clinical Finding | Condition | SNOMED |
| Open wound of lower leg | 4053604 | Clinical Finding | Condition | SNOMED |
| Osteopenia | 4195039 | Clinical Finding | Condition | SNOMED |
| Otitis externa | 380731 | Clinical Finding | Condition | SNOMED |
| Polyp of colon | 4285898 | Clinical Finding | Condition | SNOMED |
| Pressure ulcer | 135333 | Clinical Finding | Condition | SNOMED |
| Prostatism | 4016155 | Clinical Finding | Condition | SNOMED |
| Rectal hemorrhage | 4026112 | Clinical Finding | Condition | SNOMED |
| Senile hyperkeratosis | 141932 | Clinical Finding | Condition | SNOMED |
| Squamous cell carcinoma of skin | 4111921 | Clinical Finding | Condition | SNOMED |
| Traumatic wound | 46287159 | Clinical Finding | Condition | SNOMED |
| Ulcer of foot | 74719 | Clinical Finding | Condition | SNOMED |
| Ulcer of lower extremity | 197304 | Clinical Finding | Condition | SNOMED |
| Urinary incontinence | 197672 | Clinical Finding | Condition | SNOMED |
| Vaginal irritation | 4058568 | Clinical Finding | Condition | SNOMED |
| Vitamin D deficiency | 436070 | Clinical Finding | Condition | SNOMED |
| Vulval irritation | 4060207 | Clinical Finding | Condition | SNOMED |
| Wax in ear canal | 4155902 | Clinical Finding | Condition | SNOMED |

### 2. Vaccine specific analyses

#### 2.1. Index dates

**Figure S1.**
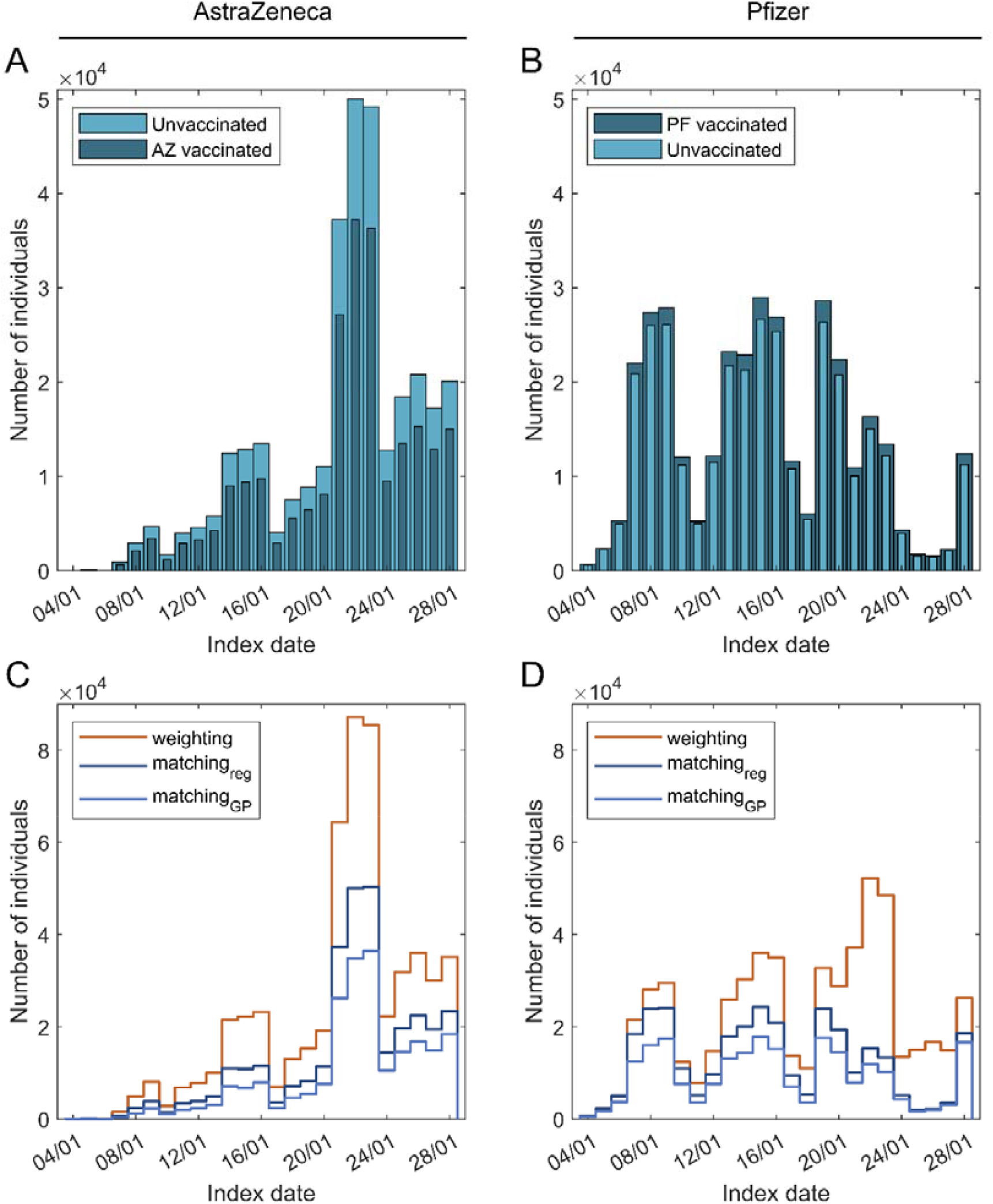
Index date distribution. (A) AstraZeneca – unvaccinated comparison. (B) Pfizer – unvaccinated comparison. (C) AstraZeneca – unvaccinated weighted and matched cohorts. (D) Pfizer – unvaccinated weighted and matched cohorts.

#### 2.2. Cohort counts

**Figure S2.**
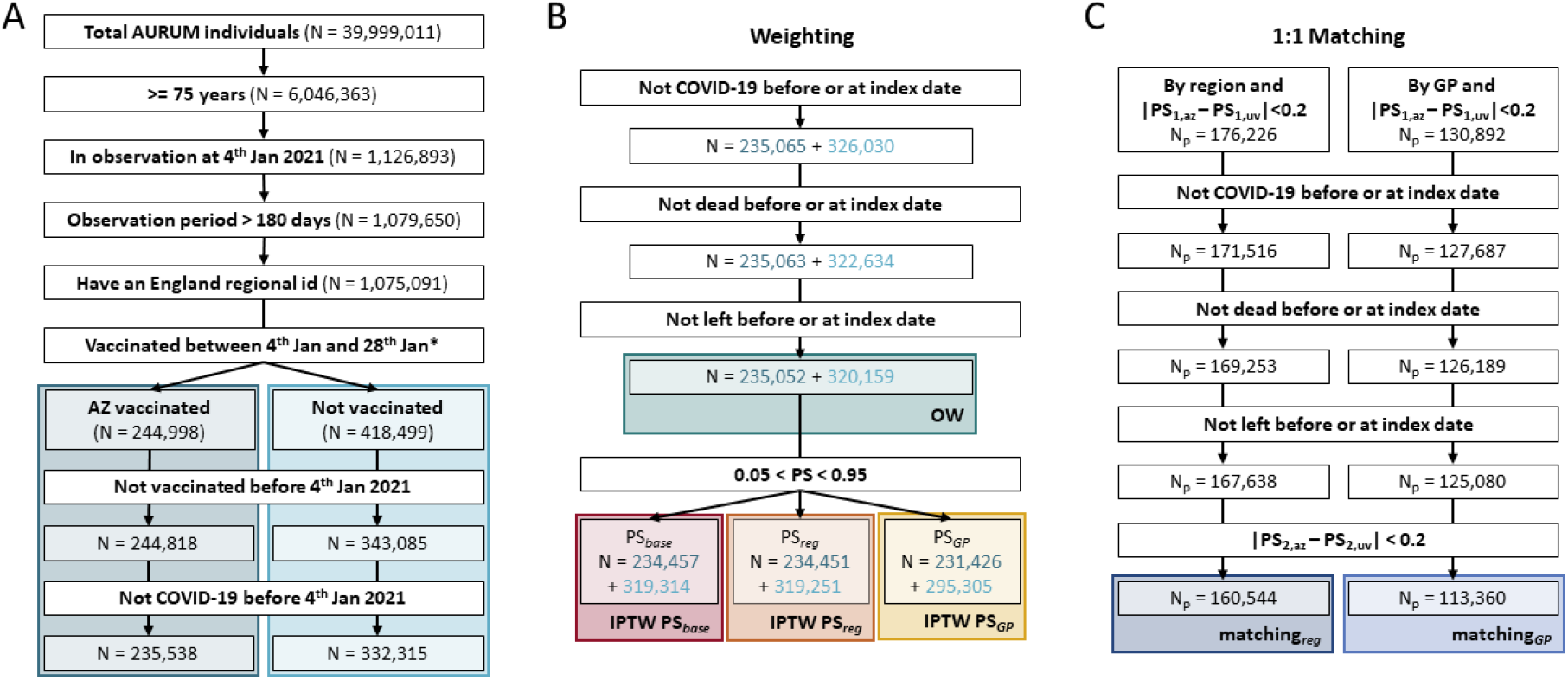
Cohorts building flowchart in AztraZeneca and unvaccinated comparison. **(A)** Flowchart to build the AstraZeneca vaccinated (AZ) and unvaccinated (UV) initial cohorts. **(B)** Flowchart to build the different weighting cohorts, the start point of these cohorts is the end of panel A. Dark blue numbers are for AZ vaccinated cohort and light for unvaccinated. **(C)** Flowchart to build the different matching cohorts, the start point of these cohorts is the end of panel A. PS_1_ and PS_2_ are the propensity scores (PS) computed at the start and index date, respectively. *At this step individuals with a record of both ChAdOx1 and BNT162b2 vaccines at the index date were excluded.

**Figure S3.**
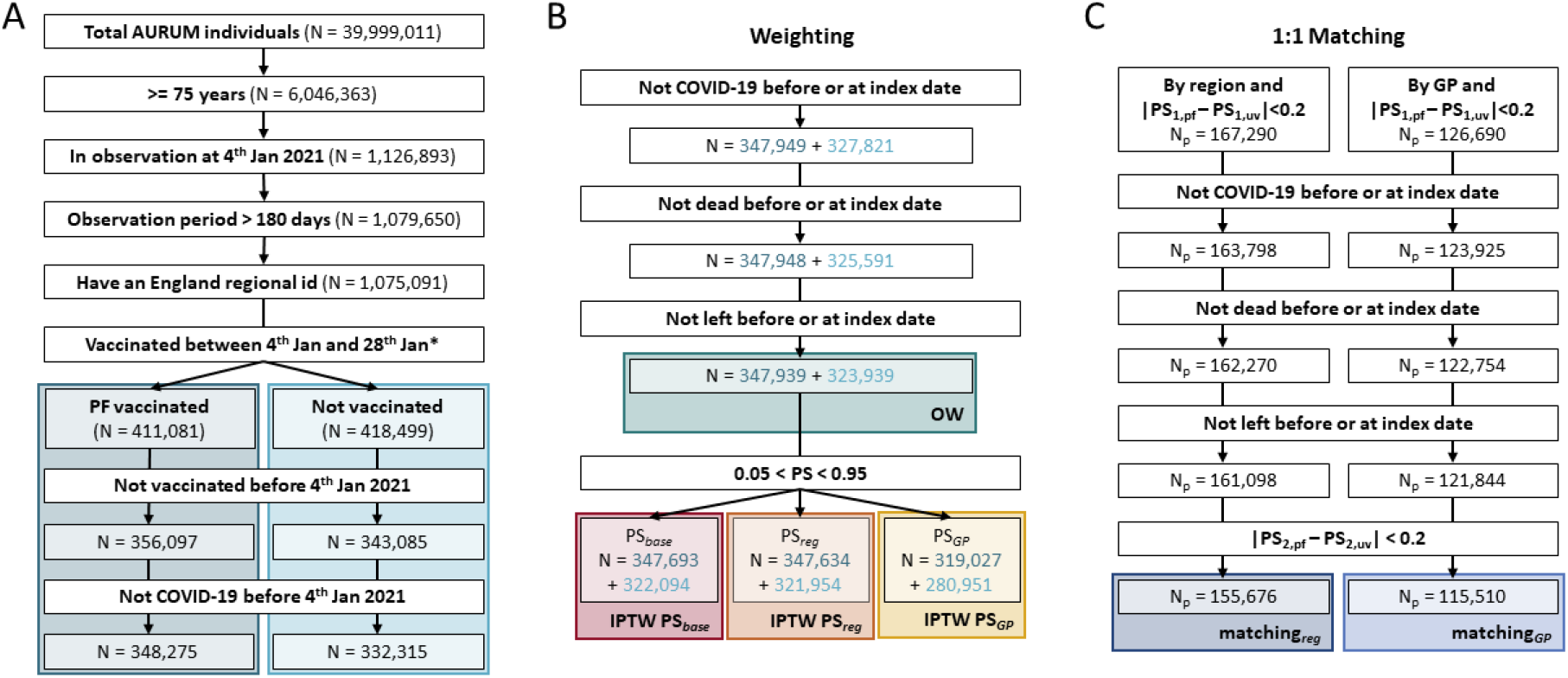
Cohorts building flowchart in Pfizer and unvaccinated comparison. **(A)** Flowchart to build the Pfizer vaccinated (PF) and unvaccinated (UV) initial cohorts. **(B)** Flowchart to build the different weighting cohorts, the start point of these cohorts is the end of panel A. Dark blue numbers are for PF vaccinated cohort and light for unvaccinated. **(C)** Flowchart to build the different matching cohorts, the start point of these cohorts is the end of panel A. PS_1_ and PS_2_ are the propensity scores (PS) computed at the start and index date, respectively. *At this step individuals with a record of both ChAdOx1 and BNT162b2 vaccines at the index date were excluded.

#### 2.3. Standardized mean differences

**Table S5.** SMDs unvaccinated vs vaccinated comparison. Only the 23 covariates with standardized mean differences greater than 0.1 are shown. 3 different periods are defined: short (−30 to -1 respect index date), mid (−180 to -31 from index date) and long (from any time prior to -181 days from index date).

| Covariate | SMD | Vaccinated mean ± STD | Unvaccinated mean ± STD |
| --- | --- | --- | --- |
| General Practice centre | 0.74 | categorical |  |
| Age group | 0.53 | categorical |  |
| Age | 0.31 | 82.2±5.4 | 79.9±5.6 |
| No response to bowel cancer screening invitation (*long*) | 0.28 | 0.19±0.39 | 0.31±0.46 |
| Normal test on bowel cancer screening programme (*long*) | 0.17 | 0.39±0.49 | 0.47±0.50 |
| Pulse rate measurement (*long*) | 0.16 | 0.88±0.32 | 0.82±0.38 |
| Diastolic blood pressure reading (*long*) | 0.15 | 1.00±0.06 | 0.98±0.14 |
| Systolic blood pressure reading (*long*) | 0.15 | 1.00±0.06 | 0.98±0.14 |
| Blood pressure reading (*long*) | 0.15 | 1.00±0.06 | 0.98±0.14 |
| Region | 0.14 | categorical |  |
| Number of visits to the GP (*long*) | 0.14 | 359±216 | 317±217 |
| Normal test on bowel cancer screening programme (*mid*) | 0.12 | 0.02±0.14 | 0.04±0.19 |
| Diastolic blood pressure reading (*mid*) | 0.12 | 0.36±0.48 | 0.30±0.46 |
| Systolic blood pressure reading (*mid*) | 0.12 | 0.36±0.48 | 0.30±0.46 |
| Blood pressure reading (*mid*) | 0.12 | 0.35±0.48 | 0.30±0.46 |
| Skin lesion (*long*) | 0.12 | 0.29±0.46 | 0.24±0.43 |
| Actinic keratosis (*long*) | 0.12 | 0.15±0.35 | 0.11±0.31 |
| Number of visits to the GP (*short*) | 0.11 | 2.28±2.38 | 1.89±2.41 |
| Wax in ear canal (*long*) | 0.11 | 0.26±0.44 | 0.21±0.41 |
| Senile hyperkeratosis (*long*) | 0.10 | 0.23±0.42 | 0.19±0.39 |
| Chronic kidney disease stage 3 (*long*) | 0.10 | 0.21±0.41 | 0.17±0.37 |
| Moderate frailty (*long*) | 0.10 | 0.16±0.37 | 0.13±0.33 |
| Basal cell carcinoma of skin (*long*) | 0.10 | 0.11±0.31 | 0.08±0.27 |

**Table S6.** SMDs AZ vaccinated vs unvaccinated comparison. Only the 25 covariates with standardized mean differences greater than 0.1 are shown. 3 different periods are defined: short (−30 to -1 respect index date), mid (−180 to -31 from index date) and long (from any time prior to -181 days from index date).

| Covariate | SMD | AZ Vaccinated mean ± STD | Unvaccinated mean ± STD |
| --- | --- | --- | --- |
| General Practice centre | 0.82 | categorical |  |
| Age group | 0.33 | categorical |  |
| Age | 0.23 | 81.7±5.7 | 79.8±5.6 |
| No response to bowel cancer screening invitation (*long*) | 0.21 | 0.22±0.42 | 0.31±0.46 |
| Region | 0.20 | categorical |  |
| Number of any covid-19 test (*mid*) | 0.18 | 0.31±1.13 | 0.08±0.55 |
| Number of PCR test (*mid*) | 0.17 | 0.23±0.89 | 0.06±0.44 |
| Pulse rate measurement (*long*) | 0.15 | 0.88±0.33 | 0.82±0.38 |
| Number of any covid-19 test (*short*) | 0.15 | 0.10±0.44 | 0.03±0.22 |
| Number of PCR test (*short*) | 0.15 | 0.10±0.43 | 0.03±0.21 |
| Diastolic blood pressure reading (*long*) | 0.14 | 1.00±0.07 | 0.98±0.14 |
| Systolic blood pressure reading (*long*) | 0.14 | 1.00±0.07 | 0.98±0.14 |
| Blood pressure reading (*long*) | 0.13 | 1.00±0.07 | 0.98±0.14 |
| Number of visits to the GP (*long*) | 0.13 | 356±219 | 317±217 |
| Number of visits to the GP (*mid*) | 0.11 | 11.8±9.2 | 10.4±9.0 |
| Sever frailty (*long*) | 0.11 | 0.09±0.28 | 0.06±0.24 |
| Skin lesion (*long*) | 0.11 | 0.29±0.45 | 0.24±0.43 |
| Number of visits to the GP (*short*) | 0.11 | 2.29±2.43 | 1.92±2.41 |
| Normal test on bowel cancer screening programme (*long*) | 0.11 | 0.02±0.15 | 0.04±0.20 |
| Blood pressure reading (*mid*) | 0.11 | 0.34±0.48 | 0.30±0.46 |
| Diastolic blood pressure reading (*mid*) | 0.10 | 0.35±0.48 | 0.30±0.46 |
| Systolic blood pressure reading (*mid*) | 0.10 | 0.35±0.48 | 0.30±0.46 |
| Moderate frailty (*long*) | 0.10 | 0.16±0.37 | 0.13±0.33 |
| Alzheimer's disease (*long*) | 0.10 | 0.04±0.20 | 0.02±0.15 |
| Bowels: fully continent (*long*) | 0.10 | 0.07±0.26 | 0.05±0.21 |

**Table S7.**
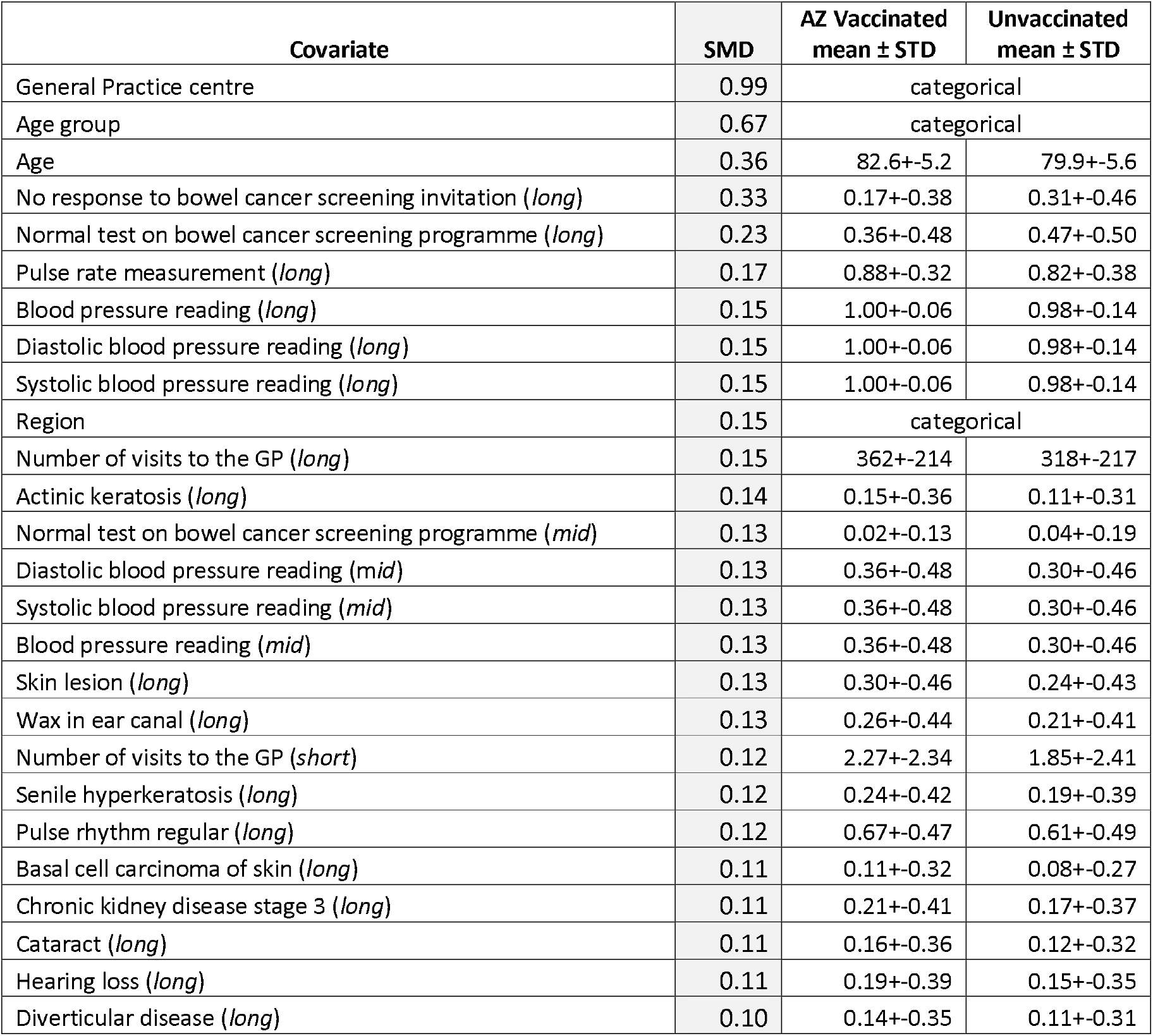
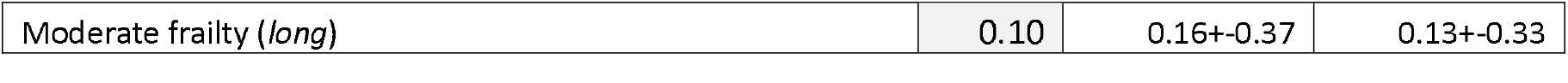
SMDs PF vaccinated vs unvaccinated comparison. Only the 27 covariates with standardized mean differences greater than 0.1 are shown. 3 different periods are defined: short (−30 to -1 respect index date), mid (−180 to -31 from index date) and long (from any time prior to -181 days from index date).

**Figure S4.**
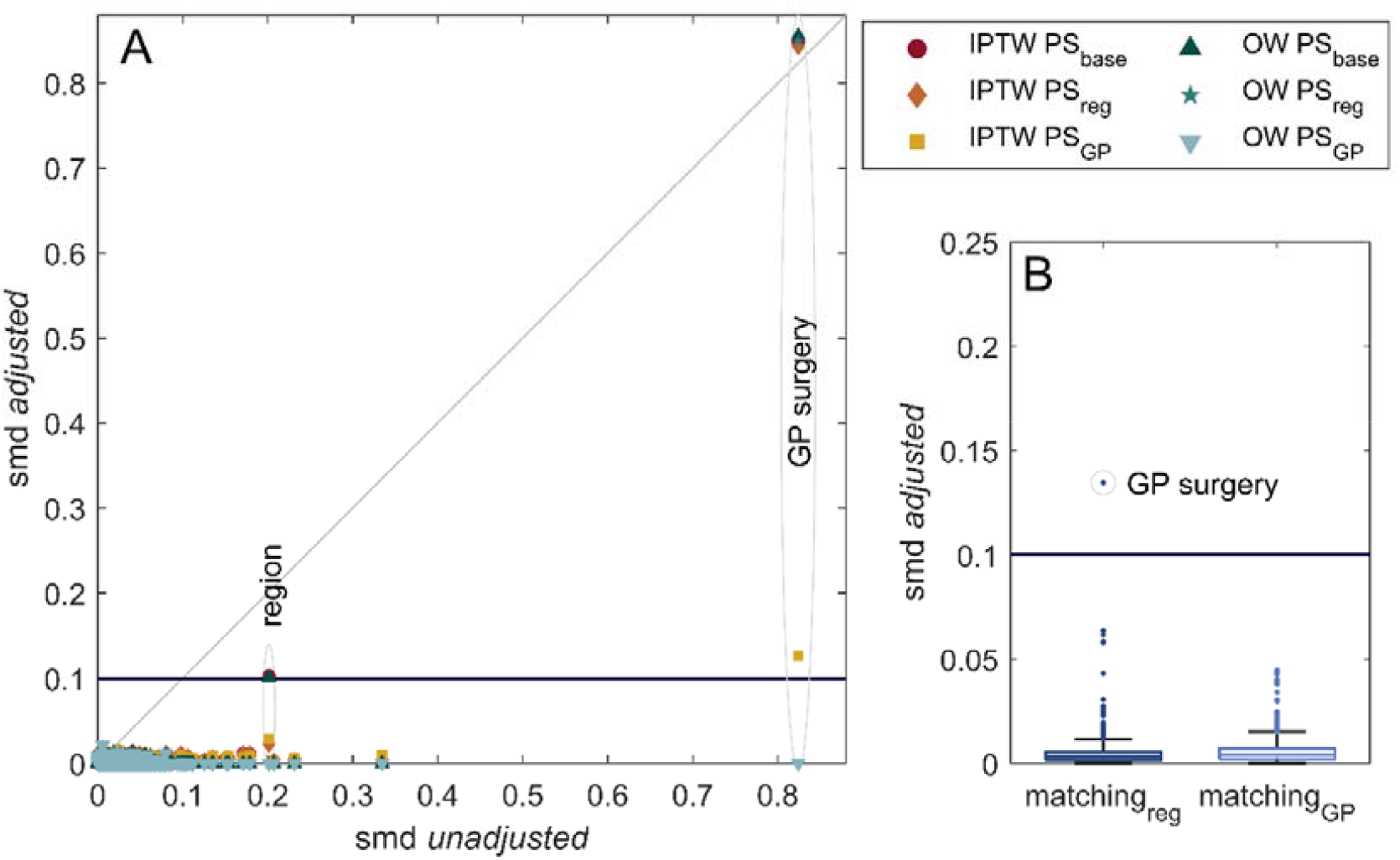
Standardized mean differences (SMD) for the different methods in AstraZeneca vaccinated and unvaccinated comparison. (A) Scatter plot to compare unadjusted smd to weighted ones. Region and GP surgery are the only covariates that remain unbalanced after some of the weightings. (B) Boxplot for the covariates smd after matching. Only GP surgery is unbalanced after regional matching.

**Figure S5.**
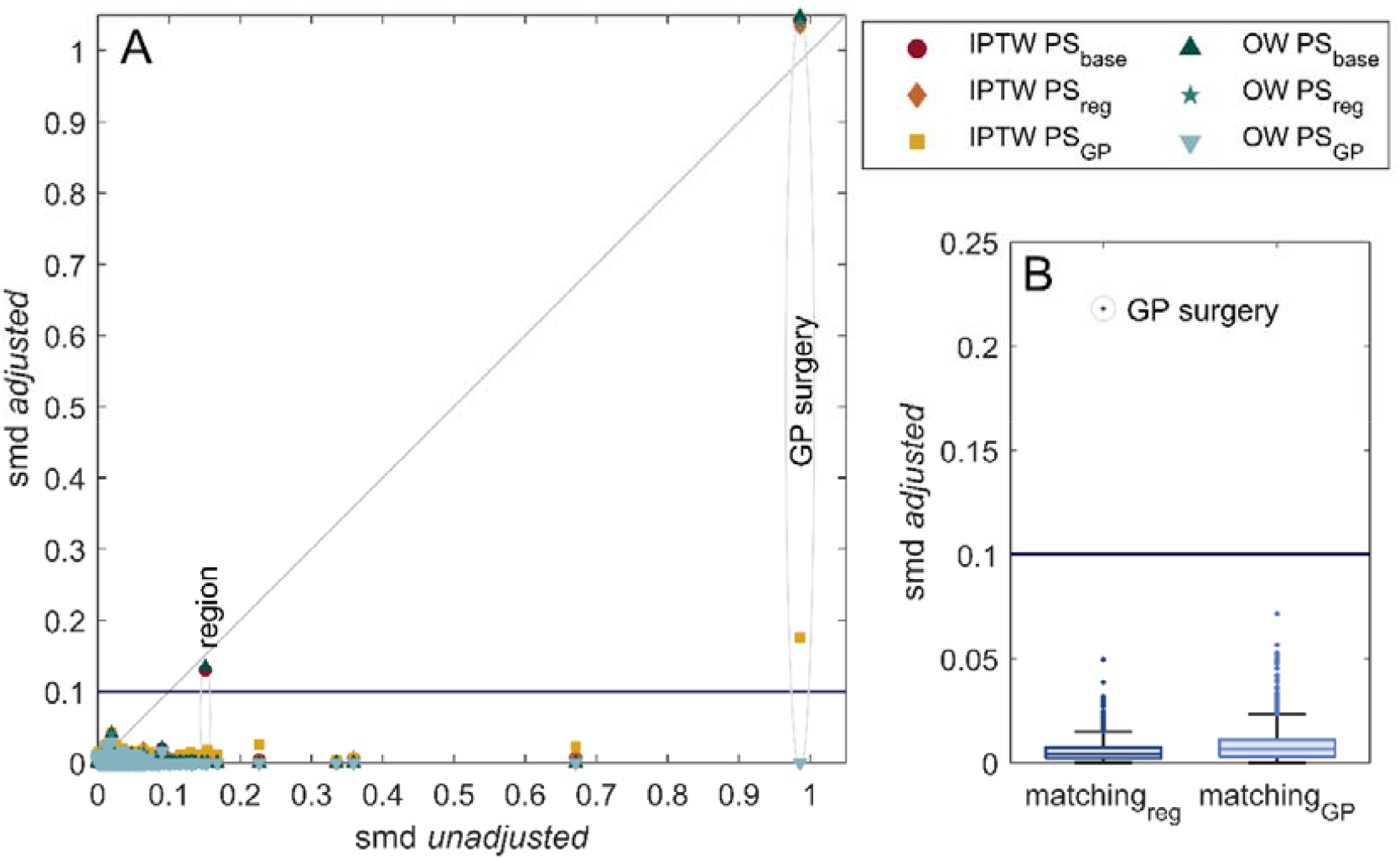
Standardized mean differences (SMD) for the different methods in Pfizer vaccinated and unvaccinated comparison. (A) Scatter plot to compare unadjusted smd to weighted ones. Region and GP surgery are the only covariates that remain unbalanced after some of the weightings. (B) Boxplot for the covariates smd after matching. Only GP surgery is unbalanced after regional matching.

#### 2.4. Outcomes distribution

**Figure S6.**
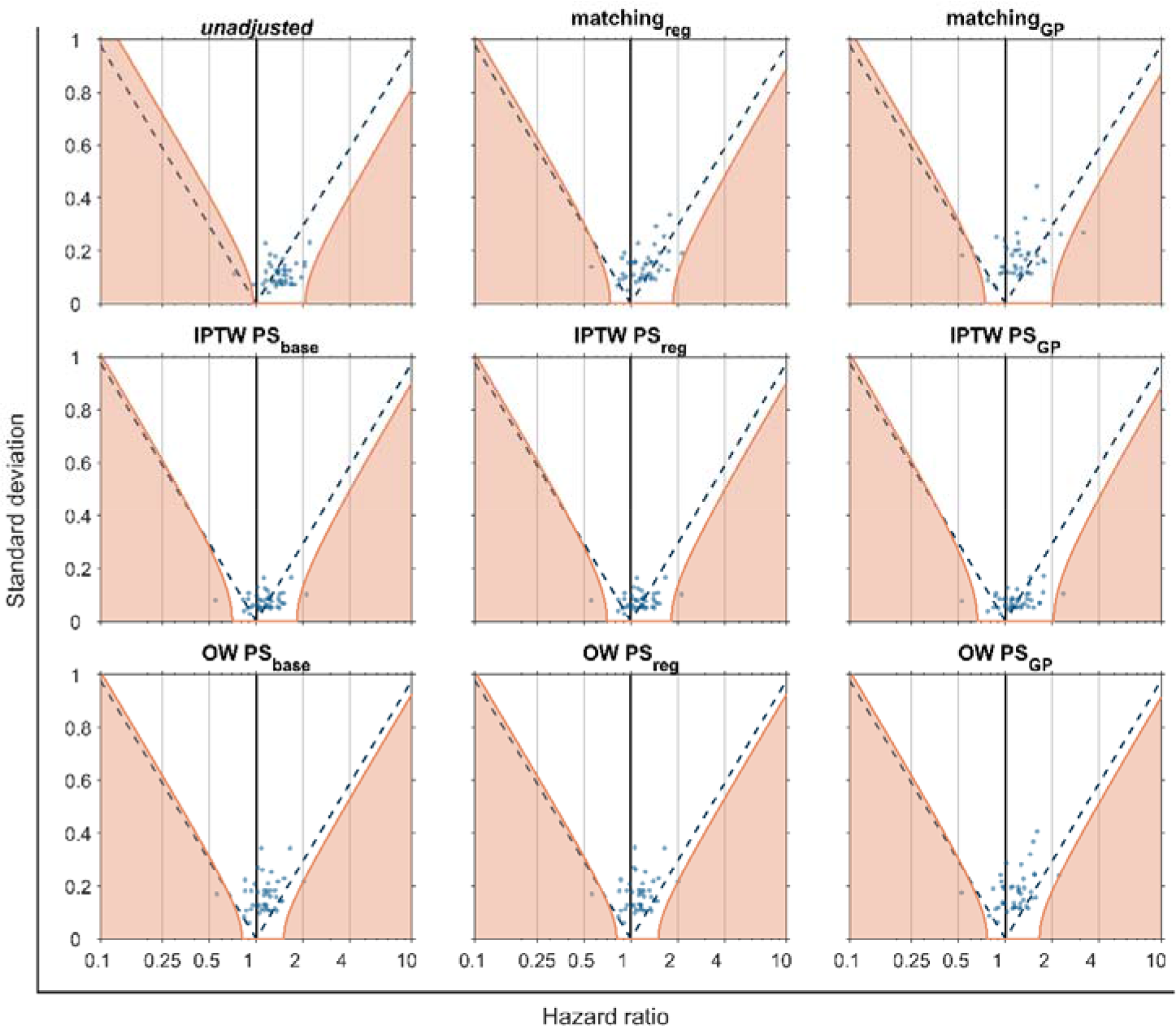
Negative control outcomes (NCO) hazard ratios and standard deviation for the AstraZeneca – unvaccinated comparison. Each blue dot represents a different NCO. Purple dashed are the significative threshold for the NCO; they are positively correlated if they are on the right of the dashed line and negatively on the left. Orange lines mark significance thresholds after calibration.

**Figure S7.**
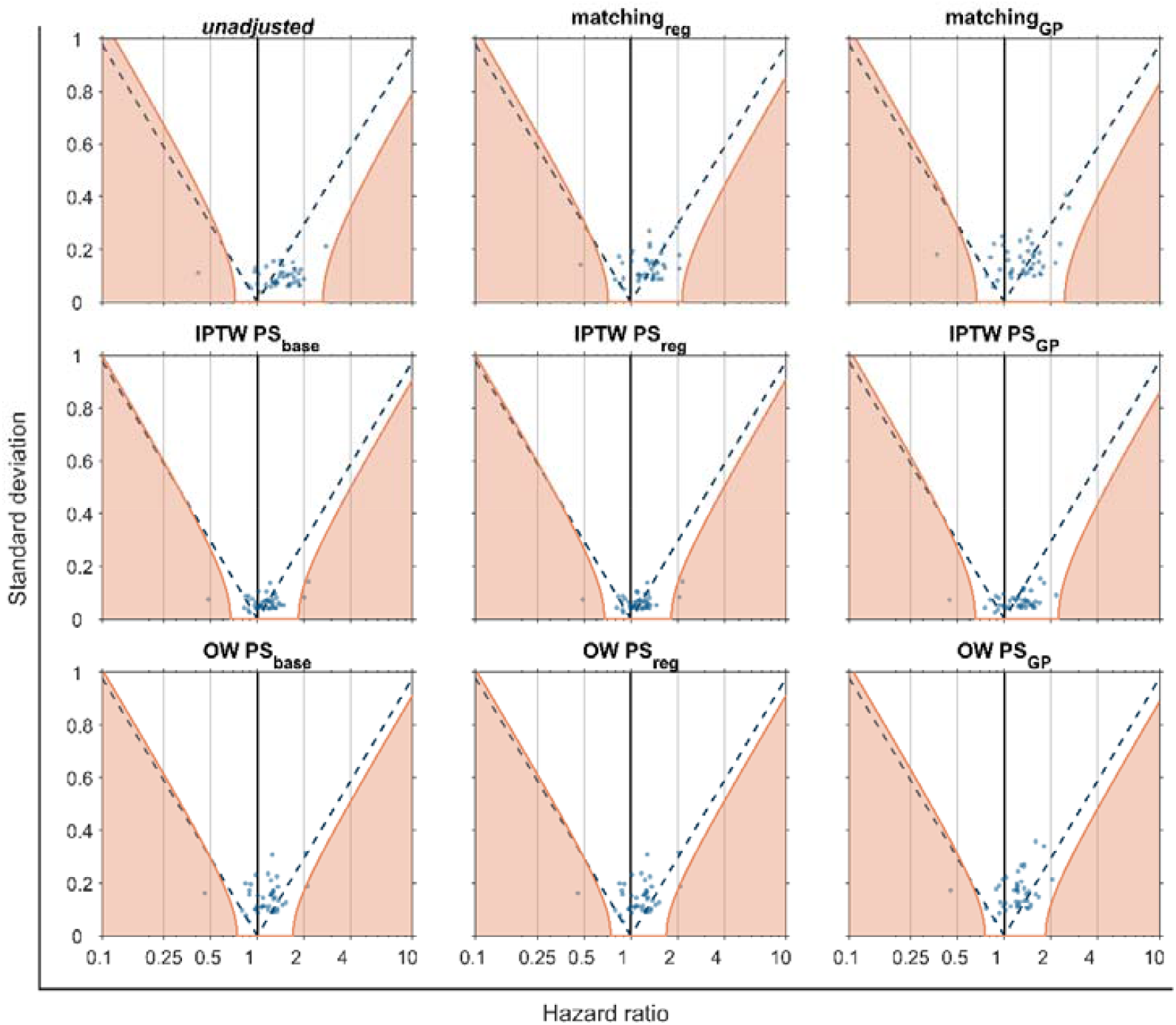
Negative control outcomes (NCO) hazard ratios and standard deviation for the Pfizer – unvaccinated comparison. Each blue dot represents a different NCO. Purple dashed are the significative threshold for the NCO; they are positively correlated if they are on the right of the dashed line and negatively on the left. Orange lines mark significance thresholds after calibration.

### 3. Hazard ratio values

**Figure S8.**
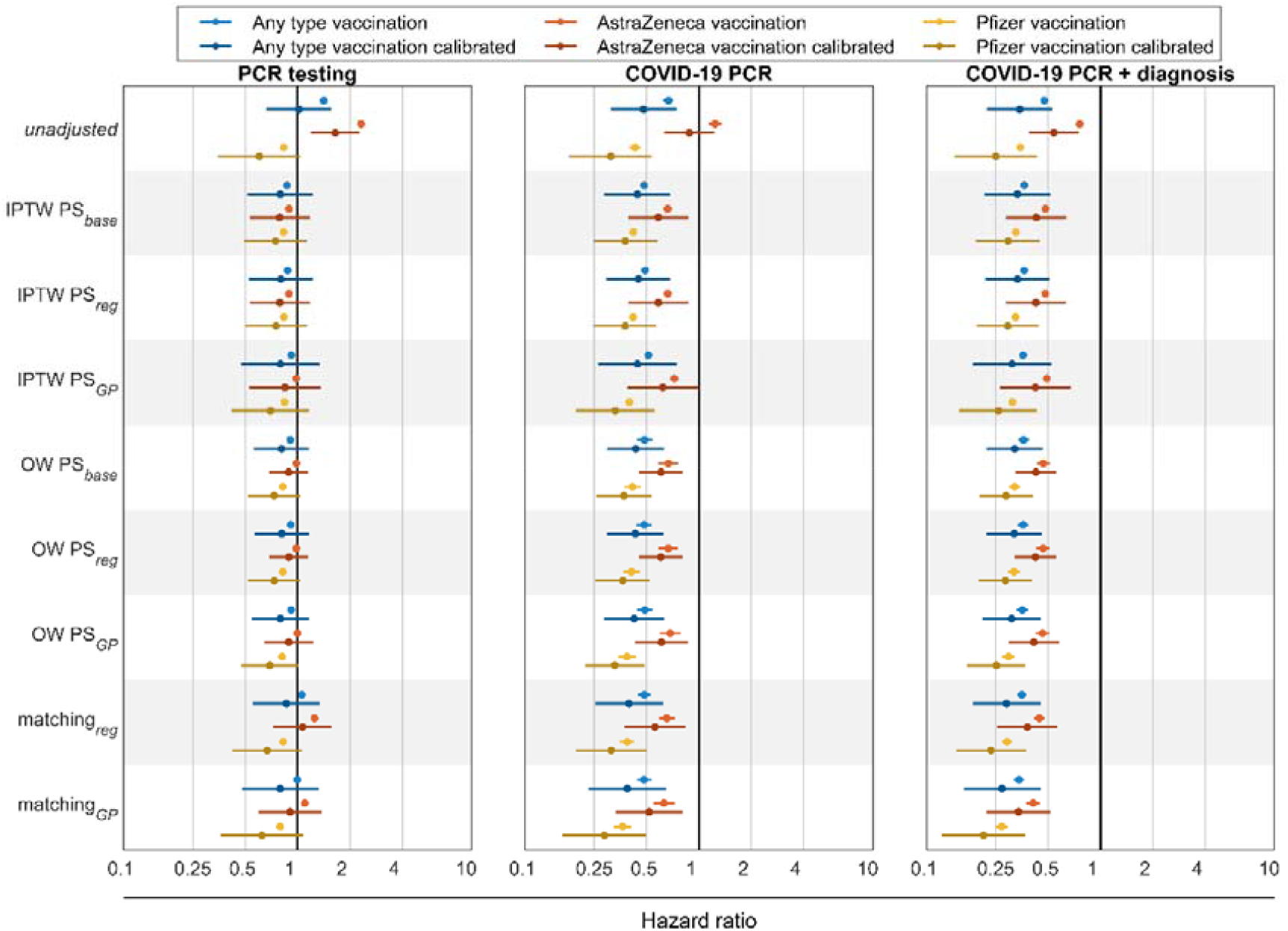
Hazard ratio for the control outcomes during all the follow-up. Each dot is the hazard ratio (HR) for a different adjustment computed with a Cox proportional hazards regression. Blue lines are for any type of vaccination compared to unvaccinated, red ones for AstraZeneca vaccinated compared to unvaccinated and yellow lines are for Pfizer vaccinated compared to unvaccinated. Darker lines are for calibrated hazard ratios. Vertical black line marks the HR = 1 threshold. In the left panel HR are for PCR testing, central and right panel for different COVID-19 definitions: only PCR positive and PCR positive or a diagnose, respectively.

**Table S8.** Hazard ratio values corresponding to figure 5. Hazard ratios were computed with a Cox proportional hazards regression censoring at day 10.

| method | calibration | Hazard ratios vaccinated |  |  |
| --- | --- | --- | --- | --- |
|  |  | Any<br>HR [95%CI] | AstraZeneca<br>HR [95%CI] | Pfizer<br>HR [95%CI] |
| PCR testing |  |  |  |  |
| unadjusted | No | 1.50 [1.44-1.55] | 2.82 [2.70-2.94] | 0.78 [0.74-0.82] |
|  | Yes | 1.08 [0.70-1.66] | 2.00 [1.45-2.77] | 0.56 [0.33-0.97] |
| IPTW PS<sub>base</sub> | No | 0.84 [0.82-0.86] | 0.94 [0.92-0.97] | 0.73 [0.71-0.76] |
|  | Yes | 0.76 [0.49-1.18] | 0.83 [0.56-1.24] | 0.66 [0.43-1.01] |
| IPTW PS<sub>reg</sub> | No | 0.84 [0.82-0.86] | 0.94 [0.92-0.97] | 0.74 [0.72-0.76] |
|  | Yes | 0.77 [0.51-1.18] | 0.83 [0.56-1.24] | 0.67 [0.44-1.01] |
| IPTW PS<sub>gp</sub> | No | 0.88 [0.86-0.90] | 1.05 [1.02-1.08] | 0.77 [0.75-0.80] |
|  | Yes | 0.76 [0.45-1.28] | 0.91 [0.57-1.45] | 0.64 [0.38-1.07] |
| OW PS<sub>base</sub> | No | 0.84 [0.80-0.90] | 1.01 [0.94-1.08] | 0.74 [0.69-0.79] |
|  | Yes | 0.75 [0.52-1.09] | 0.91 [0.70-1.19] | 0.66 [0.46-0.94] |
| OW PS<sub>reg</sub> | No | 0.85 [0.80-0.90] | 1.01 [0.94-1.08] | 0.73 [0.69-0.79] |
|  | Yes | 0.75 [0.52-1.08] | 0.91 [0.70-1.19] | 0.66 [0.46-0.93] |
| OW PS<sub>gp</sub> | No | 0.86 [0.81-0.92] | 1.02 [0.95-1.09] | 0.75 [0.70-0.81] |
|  | Yes | 0.75 [0.51-1.10] | 0.91 [0.65-1.26] | 0.64 [0.43-0.93] |
| matching<sub>reg</sub> | No | 1.05 [0.99-1.10] | 1.36 [1.28-1.44] | 0.76 [0.71-0.80] |
|  | Yes | 0.85 [0.55-1.33] | 1.16 [0.78-1.71] | 0.61 [0.39-0.97] |
| matching<sub>gp</sub> | No | 1.00 [0.94-1.06] | 1.18 [1.09-1.28] | 0.73 [0.68-0.78] |
|  | Yes | 0.80 [0.48-1.32] | 0.97 [0.64-1.49] | 0.57 [0.33-0.99] |
| **COVID-19 PCR** |  |  |  |  |
| *unadjusted* | No | 0.52 [0.48-0.57] | 1.03 [0.92-1.16] | 0.31 [0.28-0.34] |
|  | Yes | 0.38 [0.24-0.58] | 0.73 [0.52-1.03] | 0.22 [0.13-0.39] |
| IPTW PS<sub>base</sub> | No | 0.34 [0.32-0.37] | 0.53 [0.48-0.57] | 0.27 [0.25-0.29] |
|  | Yes | 0.31 [0.20-0.49] | 0.46 [0.31-0.70] | 0.25 [0.16-0.38] |
| IPTW PS<sub>reg</sub> | No | 0.35 [0.33-0.37] | 0.53 [0.49-0.57] | 0.27 [0.25-0.30] |
|  | Yes | 0.32 [0.21-0.49] | 0.47 [0.31-0.70] | 0.25 [0.16-0.38] |
| IPTW PS<sub>gp</sub> | No | 0.36 [0.34-0.38] | 0.58 [0.53-0.63] | 0.24 [0.22-0.26] |
|  | Yes | 0.31 [0.19-0.53] | 0.50 [0.31-0.80] | 0.20 [0.12-0.34] |
| OW PS<sub>base</sub> | No | 0.35 [0.30-0.41] | 0.53 [0.43-0.64] | 0.28 [0.24-0.33] |
|  | Yes | 0.31 [0.21-0.46] | 0.48 [0.35-0.66] | 0.25 [0.17-0.36] |
| OW PS<sub>reg</sub> | No | 0.35 [0.30-0.41] | 0.53 [0.44-0.64] | 0.27 [0.23-0.33] |
|  | Yes | 0.31 [0.21-0.46] | 0.48 [0.35-0.66] | 0.24 [0.17-0.36] |
| OW PS<sub>gp</sub> | No | 0.35 [0.30-0.41] | 0.55 [0.45-0.67] | 0.25 [0.21-0.30] |
|  | Yes | 0.31 [0.20-0.46] | 0.49 [0.34-0.72] | 0.21 [0.14-0.32] |
| matching<sub>reg</sub> | No | 0.36 [0.32-0.41] | 0.46 [0.40-0.54] | 0.26 [0.23-0.31] |
|  | Yes | 0.29 [0.18-0.46] | 0.40 [0.26-0.60] | 0.21 [0.13-0.34] |
| matching<sub>gp</sub> | No | 0.37 [0.32-0.43] | 0.47 [0.39-0.58] | 0.24 [0.20-0.28] |
|  | Yes | 0.30 [0.18-0.50] | 0.39 [0.25-0.62] | 0.19 [0.11-0.33] |
| **COVID-19 PCR + diagnosis** |  |  |  |  |
| *unadjusted* | No | 0.40 [0.38-0.43] | 0.70 [0.64-0.76] | 0.27 [0.25-0.29] |
|  | Yes | 0.29 [0.19-0.45] | 0.50 [0.36-0.69] | 0.20 [0.11-0.34] |
| IPTW PS<sub>base</sub> | No | 0.28 [0.27-0.30] | 0.42 [0.40-0.45] | 0.24 [0.22-0.25] |
|  | Yes | 0.26 [0.17-0.40] | 0.37 [0.25-0.56] | 0.21 [0.14-0.33] |
| IPTW PS<sub>reg</sub> | No | 0.28 [0.27-0.30] | 0.42 [0.40-0.45] | 0.24 [0.22-0.25] |
|  | Yes | 0.26 [0.17-0.39] | 0.37 [0.25-0.56] | 0.21 [0.14-0.33] |
| IPTW PS<sub>gp</sub> | No | 0.28 [0.27-0.30] | 0.44 [0.41-0.46] | 0.22 [0.21-0.24] |
|  | Yes | 0.25 [0.15-0.42] | 0.38 [0.23-0.60] | 0.19 [0.11-0.31] |
| OW PS<sub>base</sub> | No | 0.29 [0.26-0.32] | 0.40 [0.35-0.46] | 0.24 [0.21-0.27] |
|  | Yes | 0.26 [0.17-0.37] | 0.37 [0.27-0.49] | 0.21 [0.15-0.31] |
| OW PS<sub>reg</sub> | No | 0.29 [0.25-0.32] | 0.41 [0.35-0.47] | 0.24 [0.21-0.27] |
|  | Yes | 0.25 [0.17-0.37] | 0.37 [0.27-0.49] | 0.21 [0.15-0.30] |
| OW PS<sub>gp</sub> | No | 0.29 [0.25-0.32] | 0.41 [0.35-0.47] | 0.22 [0.19-0.25] |
|  | Yes | 0.25 [0.17-0.37] | 0.36 [0.26-0.52] | 0.19 [0.13-0.28] |
| matching<sub>reg</sub> | No | 0.29 [0.26-0.32] | 0.37 [0.33-0.41] | 0.23 [0.21-0.26] |
|  | Yes | 0.24 [0.15-0.37] | 0.31 [0.21-0.47] | 0.19 [0.12-0.30] |
| matching<sub>gp</sub> | No | 0.29 [0.26-0.32] | 0.37 [0.32-0.43] | 0.20 [0.18-0.23] |
|  | Yes | 0.23 [0.14-0.39] | 0.31 [0.20-0.48] | 0.16 [0.09-0.28] |

**Table S9.**
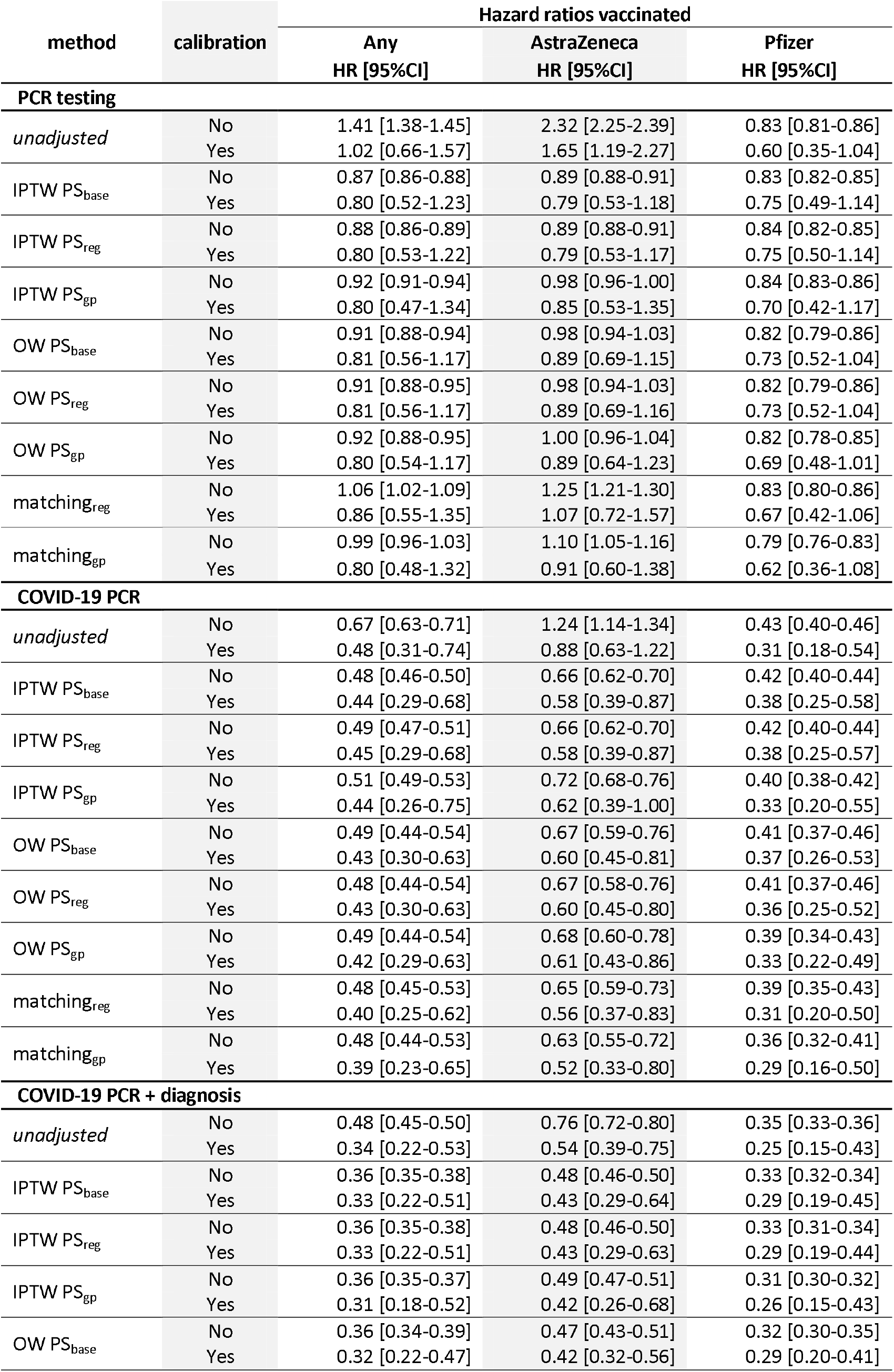
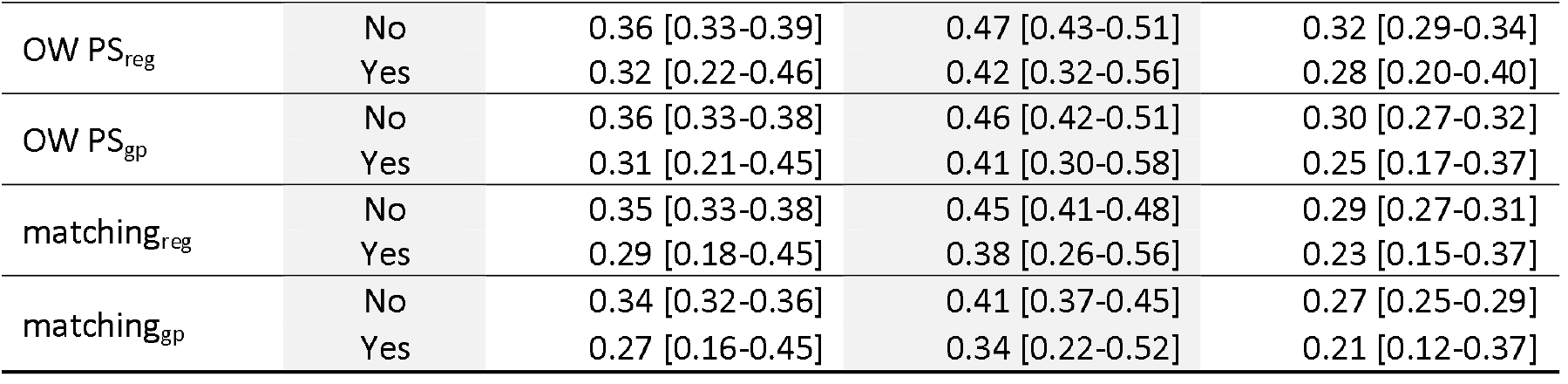
Hazard ratio values corresponding to figure S8. Hazard ratios were computed with a Cox proportional hazards regression.

**Figure S9.**
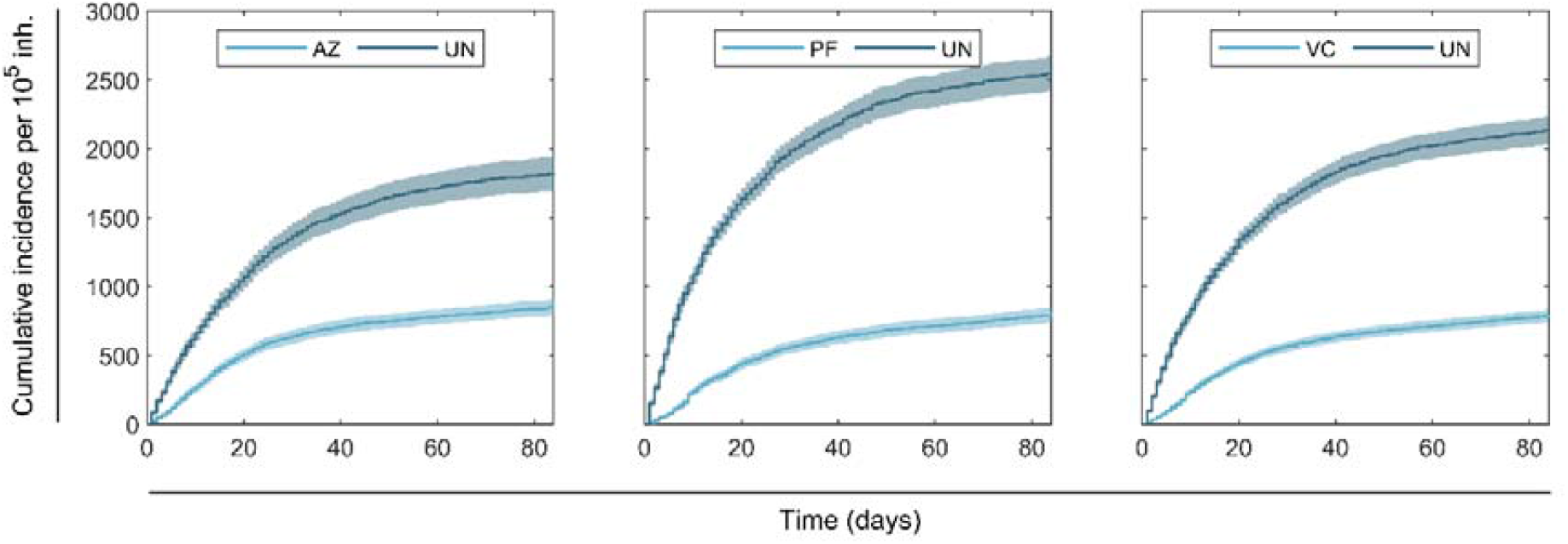
Kaplan Meier plots for COVID-19. Kaplan Meier plots using overlap weighting with PS_GP_ for COVID-19 PCR test positive or diagnose definition.

**Figure S10.**
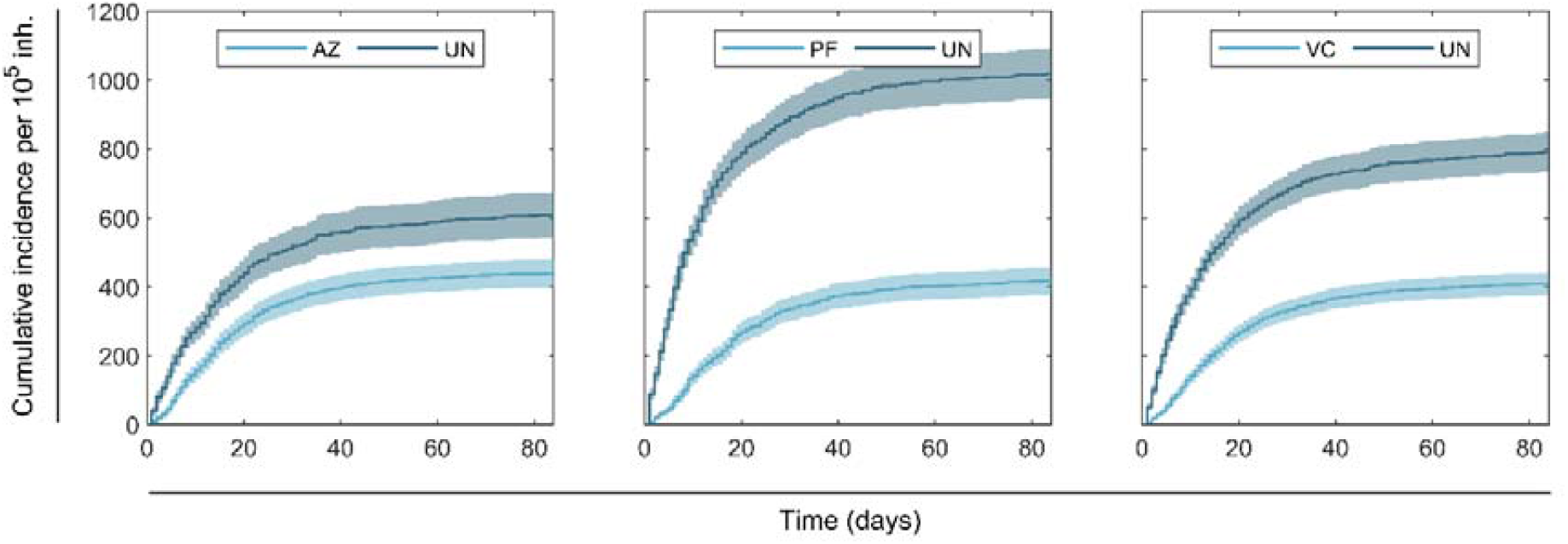
Kaplan Meier plots for COVID-19. Kaplan Meier plots using overlap weighting with PS_GP_ for COVID-19 PCR test positive definition.

## Notes

### Author Declarations

The study protocol was approved through the CPRDs Research Data Governance Process (Protocol No 21_000557).

